# An Integrated Anatomic Score for Intraprocedural Risk Stratification in Bicuspid TAVI: Development and External Validation

**DOI:** 10.64898/2026.07.18.26358381

**Authors:** Yijun Yao, Yiming Li, Tianyuan Xiong, Jianyong Wang, Weili Jiang, Yong Peng, Jiafu Wei, Sen He, Zhengang Zhao, Xin Wei, Xi Li, Wei Meng, Yuan Feng, Mao Chen

## Abstract

**Background:** Bicuspid aortic valve anatomy increases procedural complexity during transcatheter aortic valve implantation, yet outcome-oriented anatomic risk stratification for intraprocedural events remains limited.

**Aims:** We aimed to develop and externally validate an anatomy-driven score to predict a composite intraprocedural endpoint, assessed at exit from the procedure room, in bicuspid transcatheter aortic valve implantation.

**Methods:** Consecutive patients with bicuspid aortic valve undergoing transcatheter aortic valve implantation were analysed in a development cohort (N=793) and a multicentre external validation cohort (N=134). Candidate preprocedural computed tomography and echocardiographic variables were prespecified by expert consensus and refined using penalized regression with bootstrap stability selection within a domain-constrained framework. A five-indicator score (0 to 10 points) was derived from routine imaging metrics spanning the ascending aorta, aortic root, valve complex, annulus–outflow tract unit, and left ventricle, and tested using multivariable logistic regression.

**Results:** The composite intraprocedural endpoint occurred in 101/793 (12.7%) patients in the development cohort, with stepwise increases across risk strata (7.2%, 13.3%, 30.6%; p<0.001). Each 1-point increase was independently associated with higher risk (odds ratio 1.32; 95% confidence interval 1.18–1.47). A similar gradient was observed in external validation (3.1%, 10.8%, 50.0%; p=0.012; odds ratio 1.55 per point), with a C-statistic of 0.725. Higher risk categories were associated with lower early safety and higher 30-day and 1-year mortality.

**Conclusions:** An anatomy-driven score derived from routine preprocedural imaging demonstrates graded discrimination of intraprocedural risk and may inform procedural planning in bicuspid transcatheter aortic valve implantation.

## INTRODUCTION

Transcatheter aortic valve implantation (TAVI) has been established as an effective treatment for severe symptomatic aortic stenosis regardless of surgical risk.^1^ As procedural volumes have grown, so has the application of TAVI to patients with bicuspid aortic valve (BAV)—the most common congenital valvular abnormality, affecting 1–2% of the general population and accounting for up to 50% of aortic stenosis cases.^2–4^ However, BAV anatomy poses unique procedural challenges: asymmetric and extensive calcification, enlarged root dimensions, raphe-related constraints, horizontal aortic angulation, and left ventricular remodelling all contribute to higher rates of intraprocedural complications compared with tricuspid anatomy.^5,6^

Preprocedural anatomical evaluation for TAVI has accordingly expanded to span the ascending aorta, aortic root, valve complex, annulus–LVOT unit, and ventricular geometry, reflecting that prosthesis–anatomy interaction at deployment is inherently multi-level.^7^ Whether these multi-domain anatomical considerations can be translated into a quantified, outcome-anchored tool for graded intraprocedural risk assessment in the bicuspid population, where anatomical heterogeneity is most pronounced, has not been systematically addressed.

Existing tools each address only part of this gap. Generic surgical risk scores (STS, EuroSCORE) and TAVI-specific models predict mortality and comorbidity-driven outcomes but do not capture anatomy-mediated deployment risk.^8–15^ Morphologic classifications of BAV—including the Sievers, imaging-based, and 2021 International Consensus systems—standardize anatomic description but show limited correlation with procedural outcomes.^16–18^ Single-structure predictors such as calcified raphe, excess leaflet calcification, and ascending aortic dilatation have been linked to mortality, aortic injury, and paravalvular leak, yet device–anatomy interaction during TAVI occurs simultaneously across the ascending aorta, valve complex, and annulus–LVOT unit.^19,20^ An integrated, multiplanar anatomic score tailored to bicuspid TAVI is therefore lacking.

We sought to develop and externally validate BAV-Complex, an anatomy-driven score derived from routine preprocedural CT and echocardiography, to predict a composite intraprocedural endpoint assessed at exit from the procedure room in patients with BAV undergoing TAVI.

## METHODS

### Study Design

This was a two-stage development and external validation study conducted in accordance with the TRIPOD guideline. The development cohort comprised consecutive patients with CT-confirmed bicuspid aortic valve (BAV) undergoing TAVI at West China Hospital between January 2016 and June 2024. The external validation cohort was prospectively enrolled between March 2023 and October 2024 from eight centres participating in the CARRY II registry (ChiCTR2200066949).

Eligible patients had CT-confirmed BAV according to the 2021 International Consensus Classification, severe aortic valve disease deemed suitable for TAVI, and available preprocedural CT imaging. Exclusions were TAVI-in-TAVI or TAVI-in-SAVR procedures, isolated aortic regurgitation, tricuspid or quadricuspid valves, and Sievers type 2 BAV (Figure S1). Prosthesis sizing followed a supra-annular–based downsizing algorithm as previously described.^21^ Procedural details and prosthesis distribution are provided in Supplemental Method 1 and Table S1. The study was approved by the Ethics Committee of West China Hospital (2022-1003) and the institutional review boards of participating centres.

### Outcome Definitions

The primary outcome was a composite intraprocedural endpoint assessed at exit from the procedure room, adapted from the VARC-3 technical success definition: intraprocedural death, failure of device delivery, multiple transcatheter heart valves (THVs), conversion to open surgery, cardiac tamponade or aortic dissection, coronary obstruction, and haemodynamic collapse requiring resuscitation. Major vascular complications were excluded to focus on anatomy-related risk. Secondary outcomes were VARC-3 technical success, 30-day device success, 30-day early safety, and 1-year clinical efficacy (Supplemental Method 1).

### Anatomical Assessment And Score Development

All patients underwent preprocedural electrocardiogram-gated multidetector CT. Images were analysed by experienced operators and verified by a senior interventionalist (Y.F., >10 years of TAVI experience) who was blinded to intraprocedural outcomes. Left ventricular end-diastolic diameter (LVEDD) and interventricular septal thickness were obtained from preprocedural echocardiography. Acquisition protocols and measurement definitions are detailed in Supplemental Method 2.

Fifteen candidate indicators were prespecified by expert consensus and organized into five anatomical domains along the prosthesis–anatomy interaction pathway: ascending aorta, aortic root, valve complex, annulus–left ventricular outflow tract (LVOT) unit, and left ventricle (Supplemental Method 2). A prespecified, domain-constrained selection framework—combining correlation screening, univariate logistic regression, penalized regression (LASSO) with bootstrap stability assessment, and expert-guided domain filtering—was applied to derive a parsimonious five-indicator score (Supplemental Method 3; Figure S3).

### Statistical Analysis

Continuous variables are presented as median (IQR) and categorical variables as counts (percentages). Between-group comparisons used Kruskal–Wallis, chi-square, or Fisher’s exact tests as appropriate. Kaplan–Meier estimates with log-rank tests were used for 30-day and1-year mortality across risk groups.

Performance was assessed as a prespecified head-to-head comparison between a baseline STS+calcification model (calcified raphe, calcium volume, STS score) and the BAV-Complex score (0–10 points, continuous) in both cohorts. Discrimination used the C-statistic with DeLong’s test; overall performance the Brier score. Internal validation used 500-bootstrap resampling for optimism-corrected estimates (Supplemental Method 4). Independent association of the score with the composite endpoint was examined by multivariable logistic regression adjusted for age, sex, and new-generation THV.

External calibration was assessed by comparing observed and predicted event rates across the three risk strata; calibration-in-the-large and slope were estimated with a recalibration sensitivity analysis (Supplemental Method 4). Clinical utility was assessed by decision curve analysis (threshold range 0.05–0.30). Subgroup analysis stratified patients by aortic valve calcium volume (zero, 1–100 mm³, >100 mm³). Analyses were performed in R 4.3.2.

## RESULTS

### Patient characteristics

In the development cohort, 793 patients with CT-confirmed BAV were included from 2,036 consecutive TAVI procedures performed between January 2016 and June 2024, after applying the prespecified exclusion criteria. In the external validation cohort, 134 patients were enrolled from 276 screened individuals in the CARRY II registry between March 2023 and October 2024. (Figure S1)

Baseline characteristics of the development cohort are summarized in Table 1. The median age was 72 years (IQR 67–77), and 456 patients (57.5%) were male. Based on the BAV-Complex score, 276 (34.8%), 445 (56.1%), and 72 (9.1%) patients were classified into low-, intermediate-, and high-risk groups. Across increasing risk strata, male sex, chronic kidney disease, atrial fibrillation, and aortic dimensions (sinus of Valsalva perimeter, annular perimeter, and annular area) were more prevalent (all p<0.05), whereas age, body mass index, STS score, and most other comorbidities were comparable.

**Table 1.**
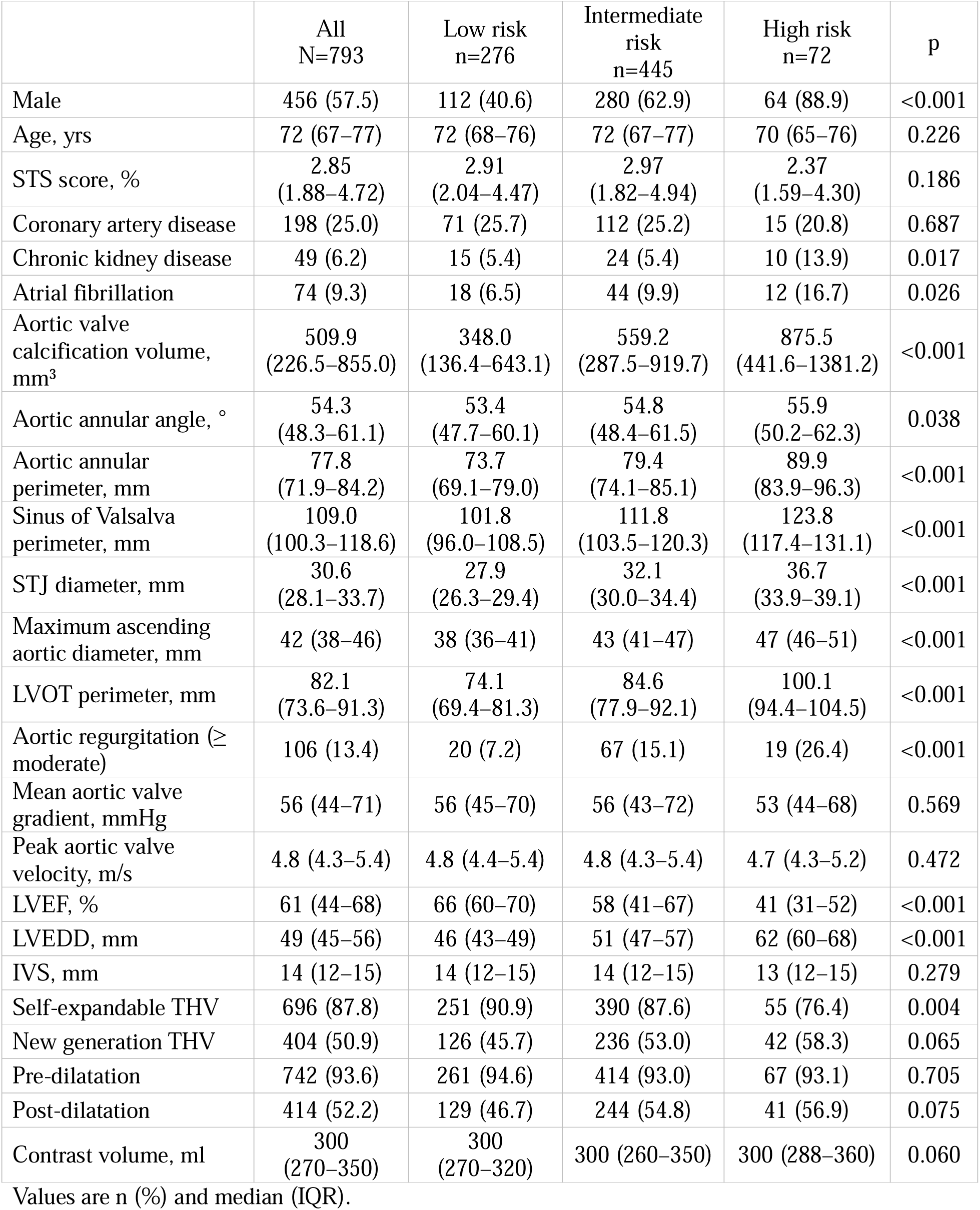
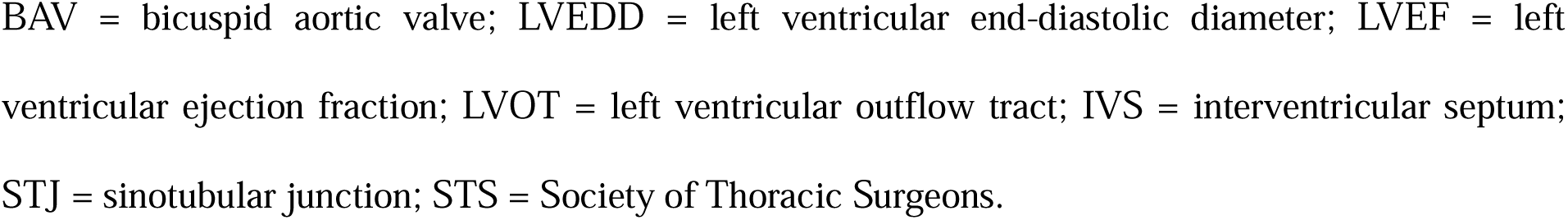
Baseline Characteristics and Procedural Details of the Development Cohort According to the BAV-Complex Score.

|  | All<br>N=793 | Low risk<br>n=276 | Intermediate<br>risk<br>n=445 | High risk<br>n=72 | p |
| --- | --- | --- | --- | --- | --- |
| Male | 456 (57.5) | 112 (40.6) | 280 (62.9) | 64 (88.9) | <0.001 |
| Age, yrs | 72 (67–77) | 72 (68–76) | 72 (67–77) | 70 (65–76) | 0.226 |
| STS score, % | 2.85<br>(1.88–4.72) | 2.91<br>(2.04–4.47) | 2.97<br>(1.82–4.94) | 2.37<br>(1.59–4.30) | 0.186 |
| Coronary artery disease | 198 (25.0) | 71 (25.7) | 112 (25.2) | 15 (20.8) | 0.687 |
| Chronic kidney disease | 49 (6.2) | 15 (5.4) | 24 (5.4) | 10 (13.9) | 0.017 |
| Atrial fibrillation | 74 (9.3) | 18 (6.5) | 44 (9.9) | 12 (16.7) | 0.026 |
| Aortic valve<br>calcification volume,<br>mm<sup>3</sup> | 509.9<br>(226.5–855.0) | 348.0<br>(136.4–643.1) | 559.2<br>(287.5–919.7) | 875.5<br>(441.6–1381.2) | <0.001 |
| Aortic annular angle, ° | 54.3<br>(48.3–61.1) | 53.4<br>(47.7–60.1) | 54.8<br>(48.4–61.5) | 55.9<br>(50.2–62.3) | 0.038 |
| Aortic annular<br>perimeter, mm | 77.8<br>(71.9–84.2) | 73.7<br>(69.1–79.0) | 79.4<br>(74.1–85.1) | 89.9<br>(83.9–96.3) | <0.001 |
| Sinus of Valsalva<br>perimeter, mm | 109.0<br>(100.3–118.6) | 101.8<br>(96.0–108.5) | 111.8<br>(103.5–120.3) | 123.8<br>(117.4–131.1) | <0.001 |
| STJ diameter, mm | 30.6<br>(28.1–33.7) | 27.9<br>(26.3–29.4) | 32.1<br>(30.0–34.4) | 36.7<br>(33.9–39.1) | <0.001 |
| Maximum ascending<br>aortic diameter, mm | 42 (38–46) | 38 (36–41) | 43 (41–47) | 47 (46–51) | <0.001 |
| LVOT perimeter, mm | 82.1<br>(73.6–91.3) | 74.1<br>(69.4–81.3) | 84.6<br>(77.9–92.1) | 100.1<br>(94.4–104.5) | <0.001 |
| Aortic regurgitation (≥<br>moderate) | 106 (13.4) | 20 (7.2) | 67 (15.1) | 19 (26.4) | <0.001 |
| Mean aortic valve<br>gradient, mmHg | 56 (44–71) | 56 (45–70) | 56 (43–72) | 53 (44–68) | 0.569 |
| Peak aortic valve<br>velocity, m/s | 4.8 (4.3–5.4) | 4.8 (4.4–5.4) | 4.8 (4.3–5.4) | 4.7 (4.3–5.2) | 0.472 |
| LVEF, % | 61 (44–68) | 66 (60–70) | 58 (41–67) | 41 (31–52) | <0.001 |
| LVEDD, mm | 49 (45–56) | 46 (43–49) | 51 (47–57) | 62 (60–68) | <0.001 |
| IVS, mm | 14 (12–15) | 14 (12–15) | 14 (12–15) | 13 (12–15) | 0.279 |
| Self-expandable THV | 696 (87.8) | 251 (90.9) | 390 (87.6) | 55 (76.4) | 0.004 |
| New generation THV | 404 (50.9) | 126 (45.7) | 236 (53.0) | 42 (58.3) | 0.065 |
| Pre-dilatation | 742 (93.6) | 261 (94.6) | 414 (93.0) | 67 (93.1) | 0.705 |
| Post-dilatation | 414 (52.2) | 129 (46.7) | 244 (54.8) | 41 (56.9) | 0.075 |
| Contrast volume, ml | 300<br>(270–350) | 300<br>(270–320) | 300 (260–350) | 300 (288–360) | 0.060 |

Values are n (%) and median (IQR).
BAV = bicuspid aortic valve; LVEDD = left ventricular end-diastolic diameter; LVEF = left ventricular ejection fraction; LVOT = left ventricular outflow tract; IVS = interventricular septum; STJ = sinotubular junction; STS = Society of Thoracic Surgeons.

Baseline characteristics of the external validation cohort are presented in Table S2. The distribution of risk categories differed from the development cohort, with 65 (48.5%), 65 (48.5%), and 4 (3.0%) patients in the low-, intermediate-, and high-risk groups, respectively.

### Development of the BAV-Complex Score

Following the prespecified selection framework (Supplemental Method 3 and Figure S3), five indicators were retained from 14 candidates across five anatomical domains: maximum ascending aortic diameter, STJ diameter, raphe length and calcification burden, LVOT–to–annular perimeter difference, and LVEDD. As shown in Figure 1, each indicator was categorized into three risk strata and assigned 0, 1, or 2 points according to clinically informed cut-offs, yielding a total score ranging from 0 to 10. Patients were stratified into low-risk (0–3 points), intermediate-risk (4–7 points), and high-risk (8–10 points) groups.

**Figure 1.**
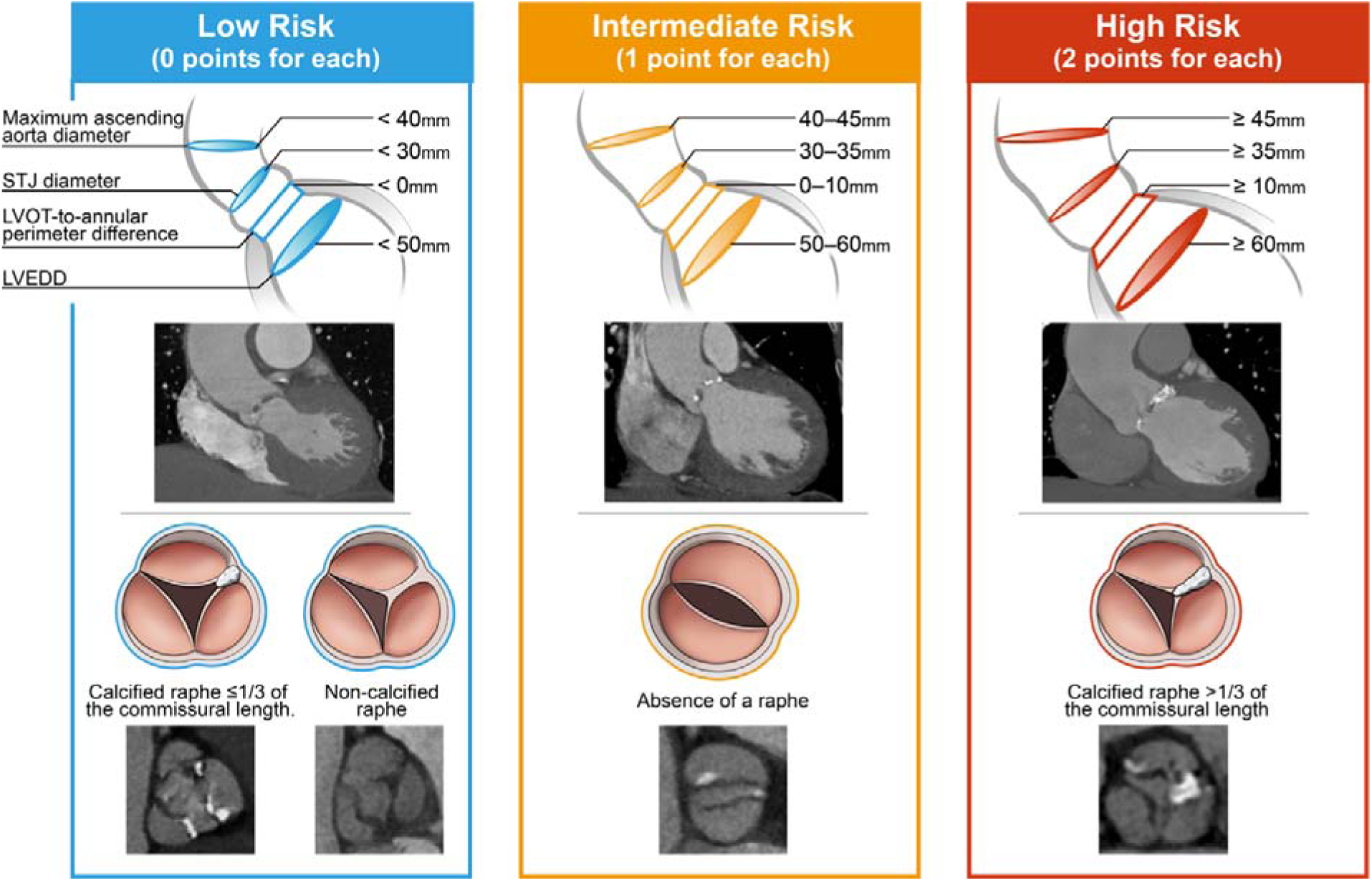
Components and Scoring Criteria of the BAV-Complex Score. The BAV-Complex score integrates five anatomical indicators spanning five device–anatomy interaction domains. Each indicator is categorized into three risk strata and assigned 0 (low risk), 1 (intermediate risk), or 2 (high risk) points as follows: maximum ascending aortic diameter (<40 mm, 40–45 mm, or ≥45 mm), sinotubular junction diameter (<30 mm, 30–35 mm, or ≥35 mm), raphe length and calcification burden (non-calcified raphe or calcified raphe involving ≤1/3 of the commissural length; absence of a raphe; or calcified raphe involving >1/3 of the commissural length), LVOT-to-annular perimeter difference (<0 mm, 0–10 mm, or ≥10 mm), and LVEDD (<50 mm, 50–60 mm, or ≥60 mm). LVEDD = left ventricular end-diastolic diameter; LVOT = left ventricular outflow tract; STJ = sinotubular junction.

### Primary outcome

The intraprocedural composite endpoint occurred in 101 of 793 patients (12.7%) in the development cohort, with stepwise increases across BAV-Complex risk categories (20/276 [7.2%], 59/445 [13.3%], and 22/72 [30.6%]; p<0.001; Figure 2 and Table 2). Detailed intraprocedural complication outcomes are reported in Table S3. A similar gradient was observed in external validation (2/65 [3.1%], 7/65 [10.8%], 2/4 [50.0%]; p=0.012). After adjustment for age, sex, and new-generation THV, each 1-point increase in BAV-Complex score was independently associated with higher risk in both cohorts (development OR 1.32, 95% CI 1.18–1.47; external OR 1.55, 95% CI 1.09–2.32; Table S4).

**Figure 2.**
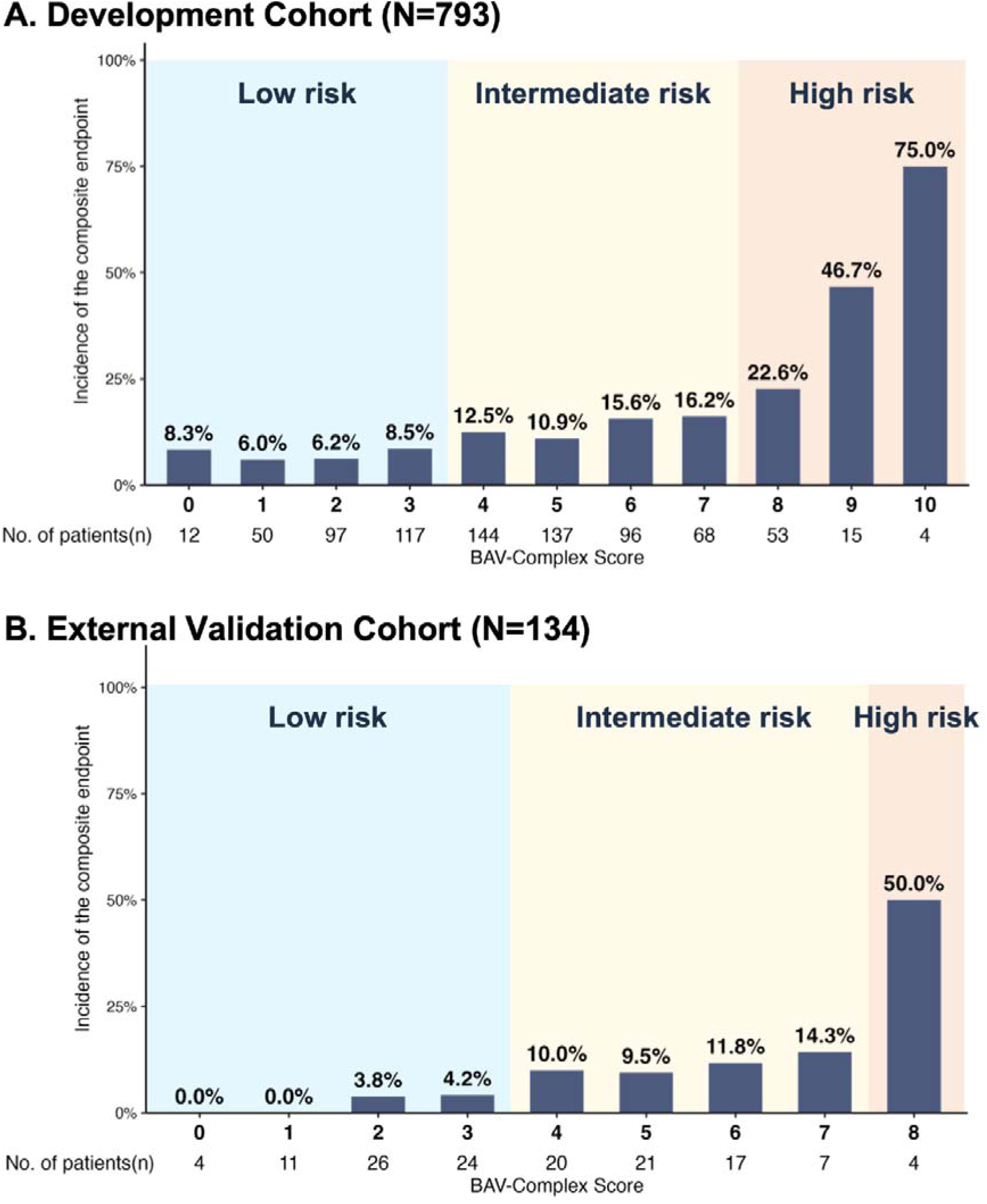
Incidence of the Composite Intraprocedural Endpoint Across BAV-Complex Score Risk Groups. Bar chart showing the incidence of the composite intraprocedural endpoint for each BAV-Complex score value (0–10) in the development cohort (**A**; N=793) and in the external validation cohort (**B**; N=134), across the three prespecified risk categories (low [0–3], intermediate [4–7], and high [8–10]). BAV = bicuspid aortic valve; TAVI = transcatheter aortic valve implantation.

**Table 2.** Procedural and Clinical Outcomes According to BAV-Complex Score Risk Groups.

| Development cohort |  |  |  |  |  |
| --- | --- | --- | --- | --- | --- |
| Variable, n (%) | All<br>N = 793 | Low risk<br>n = 276 | Intermediate risk<br>n = 445 | High risk<br>n = 72 | p |
| Composite endpoint at exit from procedure room | 101<br>(12.7) | 20<br>(7.2) | 59<br>(13.3) | 22<br>(30.6) | <0.001 |
| Technical success at exit from procedure room | 710<br>(89.5) | 258<br>(93.5) | 399<br>(89.7) | 53 (73.6) | <0.001 |
| Device success at 30 days | 559<br>(70.5) | 194<br>(70.3) | 325<br>(73.0) | 40<br>(55.6) | 0.011 |
| Early safety at 30 days | 615<br>(77.6) | 223<br>(80.8) | 349<br>(78.4) | 43<br>(59.7) | <0.001 |
| All-cause mortality at 30 days | 19<br>(2.4) | 3<br>(1.1) | 11<br>(2.5) | 5<br>(6.9) | 0.014 |
| Cardiovascular mortality at 30 days | 14<br>(1.8) | 3<br>(1.1) | 7<br>(1.6) | 4<br>(5.6) | 0.031 |
| External validation cohort |  |  |  |  |  |
|  | All<br>N = 134 | Low risk<br>n = 65 | Intermediate risk<br>n = 65 | High risk<br>n = 4 | p |
| Composite endpoint at exit from procedure room | 11 (8.2%) | 2 (3.1%) | 7 (10.8%) | 2 (50.0%) | 0.012 |
| Technical success at exit from procedure room | 124<br>(92.5%) | 62<br>(95.4%) | 60 (92.3%) | 2 (50.0%) | 0.041 |
| Device success at 30 days | 97<br>(72.4%) | 50<br>(76.9%) | 45 (69.2%) | 2 (50.0%) | 0.305 |
| Early safety at 30 days | 110<br>(82.1%) | 58<br>(89.2%) | 50 (76.9%) | 2 (50.0%) | 0.044 |
| All-cause mortality at 30 days | 2 (1.5%) | 0 (0.0%) | 1 (1.5%) | 1 (25.0%) | <0.001 |
| Cardiovascular mortality at 30 days | 2 (1.5%) | 0 (0.0%) | 1 (1.5%) | 1 (25.0%) | <0.001 |

The log-rank test was used to compare survival outcomes across BAV-Complex score risk groups at 30 days.

Bootstrap resampling (B=500) yielded an optimism-corrected C-statistic of 0.644 (optimism <0.01) and calibration slope of 1.06, indicating negligible overfitting. In the head-to-head comparison, BAV-Complex outperformed the STS+calcification model in both cohorts (development AUC 0.644 vs 0.536, DeLong p=0.003; external 0.725 vs 0.664, p=0.588), with lower Brier scores (development 0.107 vs 0.111; external 0.072 vs 0.077).

Group-level calibration in the external cohort showed modest overestimation in the low-and intermediate-risk strata (O–E, −4.2 and −2.5 percentage points) and underestimation in the high-risk stratum (O–E, +19.4 percentage points). Calibration-in-the-large was −0.315 (95% CI, −0.996 to 0.260) and the calibration slope was 1.996 (95% CI, 0.641 to 3.516), indicating a steeper risk gradient externally; recalibration improved cross-stratum agreement (Figure S4). Given the small high-risk stratum (n=4), absolute estimates should be interpreted with caution.

Decision curve analysis demonstrated consistently higher net benefit for BAV-Complex across clinically relevant thresholds in both cohorts (Figure 3). In the development cohort, net benefit differences at thresholds of 0.10 and 0.15 corresponded to approximately 1–2 additional patients per 100 appropriately stratified. In external validation, the advantage widened to 4–6 per 100 at thresholds of 0.10–0.12, where the STS+calcification model yielded negative net benefit while BAV-Complex remained positive.

**Figure 3.**
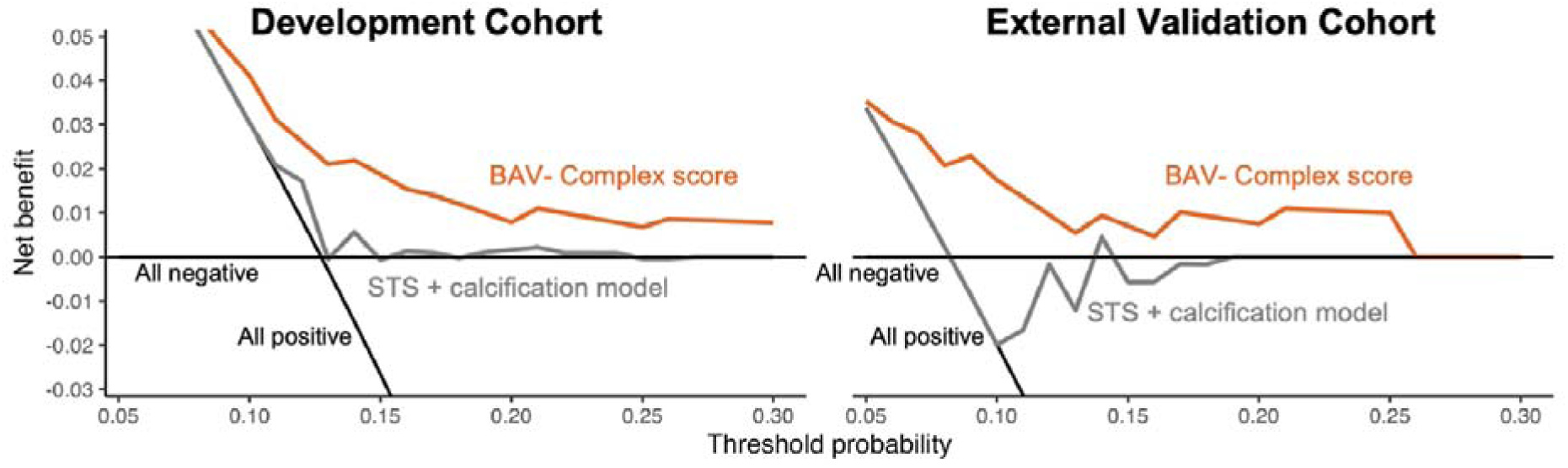
Decision Curve Analysis for the Intraprocedural Composite Endpoint. Decision curves comparing the net benefit of the BAV-Complex score model (orange) and the STS+calcification model (grey) against treat-all (black oblique line) and treat-none (black horizontal line) strategies across threshold probabilities of 0.05 to 0.30 in the development cohort **(A)** and external validation cohort **(B)**. In the development cohort, net benefit differences at thresholds of 0.10 and 0.15 corresponded to approximately 1 to 2 additional patients per 100 appropriately stratified. In external validation, BAV-Complex provided higher net benefit across thresholds of 0.05 to 0.25, with the advantage widening to 4 to 6 per 100 at thresholds of 0.10 to 0.12, where the STS+calcification model yielded negative net benefit. STS = Society of Thoracic Surgeons.

In subgroup analyses, the score retained discriminative ability among recipients of self-expandable THVs (event rates: 7.2%, 13.8%, and 34.5% across risk strata; p<0.001) and across THV generations (Tables S5–S6). Among balloon-expandable/mechanically expandable THV recipients, event rates increased numerically across risk strata (8.0%, 9.1%, and 17.6%; p=0.588), likely reflecting limited power in this smaller subgroup (n=97).

### Secondary outcomes

Across the VARC-3 secondary endpoints, higher BAV-Complex risk was consistently associated with worse procedural and short-term outcomes in the development cohort: technical success declined stepwise (93.5%, 89.7%, 73.6%; p<0.001), as did 30-day device success (70.3%, 73.0%, 55.6%; p=0.011) and early safety (80.8%, 78.4%, 59.7%; p<0.001). By 1 year, clinical efficacy did not differ significantly (93.1%, 90.6%, 86.1%; p=0.155). In the external validation cohort, similar patterns were observed for technical success (95.4%, 92.3%, and 50.0%; p=0.041) and early safety (89.2%, 76.9%, and 50.0%; p=0.044), although statistical inference was limited by the small number of high-risk patients (n=4). Differences in 30-day device success and 1-year clinical efficacy did not reach significance in this cohort (Table 2 and Table S3).

Higher BAV-Complex risk was associated with increased rates of moderate or greater paravalvular leak at 30 days (0.0%, 1.8%, 6.9%; p<0.001) and permanent pacemaker implantation (15.9%, 13.7%, 26.4%; p=0.023). All-cause mortality increased across risk strata at both 30 days (1.1%, 2.5%, 6.9%; p=0.014) and 1 year (2.5%, 5.2%, 11.1%; p=0.007) in the development cohort, with consistent gradients in external validation (30-day: 0.0%, 1.5%, 25.0%, p<0.001; 1-year: 1.5%, 3.1%, 25.0%, p=0.012). In a calcium volume-stratified analysis (zero, 1–100, >100 mm³), 12 patients (1.5%) without valve calcification had a higher 30-day pacemaker implantation rate (41.7% vs 15.2%; p=0.043) but similar 30-day mortality and composite endpoint rates (Figure S5, Table S7); this procedural pattern is consistent with the non-calcific aortic stenosis phenotype previously characterized in our centre.^22^

## DISCUSSION

We developed and externally validated the BAV-Complex score, a five-indicator (0–10 points), anatomy-driven tool derived from routine preprocedural imaging to predict a composite intraprocedural endpoint at exit from the procedure room in bicuspid TAVI. The score operationalizes the multi-domain anatomical thinking embedded in current treatment-selection frameworks^7^ into a quantified, outcome-anchored stratification tailored to patients already referred for TAVI—where prosthesis–anatomy interaction is multilevel and three-dimensional, spanning the ascending aorta, aortic root, valve complex, and the annulus–LVOT–LV unit. Across the development and multicentre validation cohorts, BAV-Complex showed graded risk stratification and aligned with procedural performance, a domain that existing comorbidity-based risk scores and descriptive BAV classifications were not designed to address. These findings support potential utility in preprocedural risk communication and procedural planning focused on anatomy-mediated deployment risk.

### Derivation Strategy and Methodologic Rationale

BAV-Complex was developed using a data-driven, domain-constrained strategy. Clinically plausible candidates prespecified by expert consensus were refined using penalized regression with bootstrap stability selection to yield a parsimonious five-component score. To mitigate overfitting, only predictors with consistent contribution across resampling iterations were retained.

### Mechanistic Interpretation of the Five Components

Each BAV-Complex component maps to a distinct “interaction zone” where prosthesis deployment can become unstable. Ascending aorta dilatation can impair coaxial delivery and device control, increasing the likelihood of malalignment during positioning and release—issues that are amplified in BAV patients who frequently exhibit horizontal root–ascending aorta remodelling.^23^ A meta-analysis confirmed that ascending aortic dilatation significantly increases the risk of aortic dissection (RR 3.55) and paravalvular regurgitation (RR 1.56) after TAVI.^24^ A dilated STJ fails to provide the necessary “upper-level” constraint, shifting the functional interaction toward a more hostile supra-annular environment. In a multicentre CT-based study of self-expandable TAVI, larger STJ diameter and height, as well as a higher STJ-to-prosthesis crown diameter ratio, were associated with a higher risk of severe prosthesis malposition.^25^ Raphe length with calcium burden captures the prototypical “rigid, asymmetric constraint” phenotype in bicuspid TAVI.

Asymmetrical calcification—particularly a calcified raphe—can impede THV sealing and expansion and is widely regarded as a key driver of procedural complexity.^26^ In the AD HOC Registry, severe annular/LVOT calcification was among the strongest predictors of moderate-to-severe paravalvular leak (aOR 5.21).^27^ Dilated left ventricles often reflect chronic volume overload, advanced myocardial remodelling, or concomitant regurgitant lesions, all of which may compromise haemodynamic tolerance during rapid ventricular pacing and valve deployment. The PARTNER 2 analysis demonstrated that larger LV end-diastolic dimensions were associated with paravalvular regurgitation and impaired reverse remodelling, partially mediating the effects of paravalvular regurgitation on mortality.^28^ Gerfer et al. demonstrated that patients with LVEF ≤40%—a condition frequently associated with LV dilation—experienced intraprocedural cardiopulmonary resuscitation in 17% of cases versus 5.8% in those with preserved LVEF (p<0.001), with intraprocedural defibrillation required in 13% versus 5.4% (p=0.002).^29^

### Clinical Implications

The moderate discrimination (AUC 0.644) reflects the intrinsic difficulty of predicting intraprocedural events from static preprocedural imaging. Nonetheless, BAV-Complex significantly outperformed the STS+calcification model (p=0.003) and yielded consistently higher net benefit on decision curve analysis, corresponding to 1–2 additional patients per 100 appropriately stratified in the development cohort and 4–6 per 100 in external validation, where the comparator fell below the treat-none reference.

This stepwise risk gradient was consistent on external validation and persisted after multivariable adjustment, with the high-risk stratum showing an approximately fourfold higher event rate than the low-risk stratum and paralleling reductions in technical success and early safety. These findings support use of BAV-Complex preprocedurally to inform heart-team discussions, anticipate deployment complexity, and tailor procedural preparation.

### Study Limitations

Several limitations should be acknowledged. First, the score does not include supra-annular parameters, for which measurement remains non-standardized. Second, CT measurements were obtained by manual analysis; inter-observer variability cannot be excluded despite expert cross-checking. Third, the high-risk stratum in the external cohort comprised only four patients, limiting stratum-specific precision; nonetheless, continuous-variable analysis using all 134 patients (OR 1.55 per point; p=0.021) supported discriminative ability. Ongoing CARRY II enrolment will permit further validation.

## CONCLUSIONS

In patients with BAV undergoing TAVI, we developed and externally validated the BAV-Complex score, a five-indicator (0–10 points) anatomy-driven tool derived from routine preprocedural imaging to predict intraprocedural endpoints at procedure completion. By translating multi-domain anatomical evaluation into a quantified, transportable risk gradient, the score may complement existing clinical risk models and inform preprocedural discussions and risk-tailored planning for anatomy-mediated deployment risk in bicuspid TAVI.

### Impact on Daily Practice

The BAV-Complex score (0–10 points), built from five routine preprocedural imaging indicators, provides graded intraprocedural risk stratification in bicuspid TAVI that outperforms a STS plus calcification model and shows consistent risk ordering on multicentre external validation. By integrating ascending aortic dilatation, sinotubular junction width, raphe length and calcification burden, LVOT-to-annular geometry, and left ventricular dimension, the score may be applied at the time of preprocedural CT review to inform heart-team discussions, anticipate deployment complexity, and tailor procedural preparation in bicuspid anatomy referred for TAVI.

## Data Availability

The de-identified data supporting the findings of this study are available from the corresponding author upon reasonable request.

## Acknowledgments

The authors thank Dan Liu, Youfang Huang (Department of Cardiology, West China Hospital, Sichuan University) and the whole follow-up team for their contributions to patient follow-up.

## Ethics Approval

The study was conducted in accordance with the Declaration of Helsinki. Ethical approval was obtained from the Ethics Committee of West China Hospital (approval number 2022-1003) and the institutional review boards of all participating centres of the CARRY II registry (ChiCTR2200066949). For patients whose data were analysed retrospectively prior to the 2022 ethics approval, the requirement for written informed consent was waived by the ethics committee given the retrospective observational design and the use of de-identified data. All patients enrolled prospectively thereafter provided written informed consent prior to enrolment.

## Abbreviations

AS: aortic stenosis
AUC: area under the curve
BAV: bicuspid aortic valve
CI: confidence interval
CT: computed tomography
IQR: interquartile range
LVEDD: left ventricular end-diastolic diameter
LVOT: left ventricular outflow tract
OR: odds ratio
PVL: paravalvular leak
STJ: sinotubular junction
STS: Society of Thoracic Surgeons
TAVI: transcatheter aortic valve implantation
THV: transcatheter heart valve
VARC-3: Valve Academic Research Consortium 3

## Supplemental materials

**Supplemental Method 1.** Procedural and Follow-up Considerations Specific to Bicuspid TAVI **Supplemental Method 2.** Imaging Acquisition, Measurement Definitions, and Candidate Indicators **Supplemental Method 3.** Variable Selection and Score Construction

**Supplemental Method 4.** Internal Validation, External Calibration, and Missing Data

**Table S1.** Features and Cohort Distribution of TAVI Prostheses.

**Table S2.** Baseline Characteristics and Procedural Details of the Multicentre External Validation Cohort According to the BAV-Complex Score.

**Table S3.** Procedural and Clinical Outcomes According to BAV-Complex Score Risk Groups.

**Table S4.** Multivariable Analysis of the Composite Endpoint in the Development Cohort and External validation cohort

**Table S5.** Outcomes between Self-expandable and Balloon-expandable THVs in Development Cohort.

**Table S6.** Outcomes between First-generation and New-generation THVs in Development Cohort.

**Table S7.** Baseline, Anatomic, and Procedural Characteristics of Patients without Aortic Valve Calcification (n=12) versus the Rest of the Development Cohort (n=781).

**Figure S1.** Study Flowchart.

**Figure S2.** Incidence of the Composite Endpoint by Individual Anatomical Parameters in the Development Cohort.

**Figure S3.** Anatomical variable screening, stability assessment, and contribution analysis for model development.

**Figure S4.** Group-Level External Calibration Before and After Recalibration.

**Figure S5.** Procedural and Outcome Comparison Across Aortic Valve Calcium Volume Strata.

