## Supplemental Material for "An Integrated Anatomic Score for Intraprocedural Risk Stratification in Bicuspid TAVI: Development and External Validation"

### Supplemental materials

**Supplemental Method 1.** Procedural and Follow-up Considerations Specific to Bicuspid TAVI

**Supplemental Method 2.** Imaging Acquisition, Measurement Definitions, and Candidate Indicators

**Supplemental Method 3.** Variable Selection and Score Construction

**Supplemental Method 4.** Internal Validation, External Calibration, and Missing Data

**Table S1.** Features and Cohort Distribution of TAVI Prostheses.

**Table S2.** Baseline Characteristics and Procedural Details of the Multicentre External Validation Cohort According to the BAV-Complex Score.

**Table S3.** Procedural and Clinical Outcomes According to BAV-Complex Score Risk Groups.

**Table S4.** Multivariable Analysis of the Composite Endpoint in the Development Cohort and External validation cohort

**Table S5.** Outcomes between Self-expandable and Balloon-expandable THVs in Development Cohort.

**Table S6.** Outcomes between First-generation and New-generation THVs in Development Cohort.

**Table S7.** Baseline, Anatomic, and Procedural Characteristics of Patients without Aortic Valve Calcification (n=12) versus the Rest of the Development Cohort (n=781).

**Figure S1.** Study Flowchart.

**Figure S2.** Incidence of the Composite Endpoint by Individual Anatomical Parameters in the Development Cohort.

**Figure S3.** Anatomical variable screening, stability assessment, and contribution analysis for model development.

**Figure S4.** Group-Level External Calibration Before and After Recalibration.

**Figure S5.** Procedural and Outcome Comparison Across Aortic Valve Calcium Volume Strata.

### Supplemental Method 1. Procedural and Follow-up Considerations Specific to Bicuspid TAVI

All procedures were performed in a hybrid catheterisation laboratory under fluoroscopic guidance, with local anaesthesia and conscious sedation as the default approach. Transfemoral access was used whenever feasible, with ultrasound-guided puncture and pre-close suture-based closure (Perclose ProGlide, Abbott Vascular). A temporary pacing lead was placed via the contralateral femoral vein or internal jugular vein.

Self-expanding THVs were preferred given their adaptability to the elliptical and heavily calcified geometry typical of BAV anatomy; prosthesis distribution across both cohorts is summarised in Table S1. Routine balloon pre-dilation was performed with undersized balloons. A snare-assisted delivery technique was used in cases of horizontal aorta (annular angulation ≥50°) or dilated ascending aorta (≥45 mm). Valve deployment was performed under rapid ventricular pacing (180–200 bpm). In patients with anticipated coronary obstruction risk on preprocedural CT (low coronary ostium height, shallow sinus of Valsalva, or bulky leaflet calcification), coronary protection with pre-positioned wires or undeployed stents was applied. Post-dilation, percutaneous paravalvular leak closure, and implantation of a second THV were performed selectively as indicated. Dual antiplatelet therapy was initiated in the absence of oral anticoagulation and exceptionally high bleeding risk.

Clinical follow-up was scheduled at 30 days, 3 months, 6 months, and 12 months, conducted by heart-team members and trained research staff through outpatient visits, telephone, or video consultation when in-person visits were not feasible.

### Supplemental Method 2. Imaging Acquisition, Measurement Definitions, and Candidate Indicators

Preprocedural electrocardiogram-gated multidetector CT was performed with a second-generation dual-source system (SOMATOM Definition Flash, Siemens) or a spectral CT system (Revolution, GE Healthcare), using approximately 80 mL of intravenous iodinated contrast, with systolic-phase reconstruction (25–35% R-R interval). Images were analysed in OsiriX (OsiriX Foundation) by two experienced physicians and verified by Y.F.

All structural measurements were obtained in the systolic phase. BAV was classified per the 2021 International Consensus Classification, with the partial-fusion phenotype considered BAV. Raphe length was expressed as the proportion of raphe to the total commissural edge; aortic valve calcium volume was quantified on contrast-enhanced images at a threshold of 850 HU; the LVOT was measured 4 mm below the annular plane; the maximum ascending aortic diameter was taken at the widest cross-section along the centreline; and annular angulation was defined as the angle between the annular plane and the CT coronal plane.

Fifteen candidate indicators were prespecified by a multidisciplinary panel of interventionalists (each >10 years of TAVI experience), organised into five anatomical domains: ascending aorta (maximum ascending aortic diameter); aortic root (sinotubular junction [STJ] diameter, sinus of Valsalva perimeter, left and right coronary ostium heights); aortic valve (calcification volume; raphe length and calcification burden, categorised as low-risk [non-calcified raphe or calcified raphe ≤1/3 of the commissural length], intermediate-risk [absence of raphe], or high-risk [calcified raphe >1/3 of the commissural length]); annulus–LVOT complex (annular perimeter, annular area, annular angulation, LVOT perimeter, and LVOT-to-annular perimeter difference); and left ventricle (LVEDD, interventricular septal thickness, and LVEF from echocardiography). The final score was required to include at least one indicator per domain to preserve three-dimensional anatomical representation.

### Supplemental Method 3. Variable Selection and Score Construction

Variable selection followed a prespecified, domain-constrained framework combining data-driven and expert criteria (Figure S3). Regression analyses used complete-case data; Spearman correlations used pairwise complete observations.

**Step 1 — Correlation screening.** Spearman correlations were computed across all 14 candidates; pairs with |r| ≥ 0.70 were flagged as redundant. For mathematically interdependent metrics (annular perimeter, LVOT perimeter, and their difference), selection was guided by monotonicity of event rates across ordered strata (Figure S2). LVOT perimeter was excluded; annular perimeter and LVOT-to-annular perimeter difference were retained.

**Step 2 — Univariate association.** Each remaining candidate was assessed by univariate logistic regression with the composite endpoint as the dependent variable, and discrimination quantified by AUC. Continuous predictors were standardised to clinically meaningful increments (per-unit scaling detailed in Figure S3B).

**Step 3 — Penalised regression with stability selection.** LASSO logistic regression was fitted with λ tuned by 10-fold cross-validation (AUC metric). Stability was assessed by 200 bootstrap resamples; variables with non-zero coefficients at λ₁SE in ≥50% of samples were considered stable.

**Step 4 — Domain-constrained final selection.** Candidates were prioritised within each domain by the product of univariate AUC and bootstrap selection frequency, with ties broken by clinical relevance and measurement reliability. At least one variable per anatomical domain was retained.

**Step 5 — Leave-one-out contribution analysis.** The incremental discrimination of each retained variable was quantified by the reduction in AUC upon its removal from the full multivariable model, expressed as a percentage of the total AUC gain beyond 0.5.

Each retained variable was categorised into three risk strata and assigned 0, 1, or 2 points according to clinically informed cut-offs (Figure 1), yielding a total score of 0–10. Patients were stratified as low-risk (0–3), intermediate-risk (4–7), or high-risk (8–10).

### Supplemental Method 4. Internal Validation, External Calibration, and Missing Data

**Internal validation.** Bootstrap resampling (B = 500) was used to quantify optimism in the development cohort. In each iteration, the logistic model (outcome ~ BAV-Complex score) was refitted on a resampled dataset, and performance (C-statistic and calibration slope) was evaluated on both the bootstrap and original samples. Optimism-corrected metrics were obtained by subtracting mean optimism from apparent performance.

**External calibration.** A logistic model with the three prespecified risk strata as predictors (outcome ~ risk group) was fitted in the development cohort, and predicted probabilities were applied to the external cohort without refitting. Group-level calibration was summarised by comparing observed event rates with mean predicted probabilities within each stratum, reported as observed-minus-predicted differences in absolute percentage points and visualised on a 45° calibration plot. Calibration-in-the-large was estimated by an offset logistic model, and the calibration slope by regressing the outcome on the linear predictor from the development model. Predicted probabilities were truncated away from 0 and 1 before logit transformation for numerical stability.

**Recalibration sensitivity analysis.** Recalibrated probabilities were generated in the external cohort from the estimated calibration intercept and slope and displayed alongside the original predictions (Figure S4).

**Missing data.** Missingness for demographics, comorbidities, and echocardiographic parameters was <10%, with no patient missing all CT or echocardiographic data. Missing demographic and continuous echocardiographic values were imputed by cohort median. Missing aortic or mitral regurgitation grades were assumed to be less than moderate.

### Table S1. Features and Cohort Distribution of TAVI Prostheses.

|  | **Prosthesis images** | **Development Cohort** | **External Validation Cohort** | **Expansion Mechanism** | **Recapturability** | **Outer Sealing Skirt** | **New-generation** |
| --- | --- | --- | --- | --- | --- | --- | --- |
| VenusA | 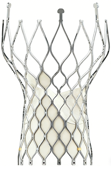 | 341 (43.0) | 1 (0.7) | SE | None | No | No |
| VenusA-Plus |  | 125 (15.8) | 127 (94.8) | SE | Partial | No | Yes |
| VenusA-Pro |  | 0 (0) | 6 (4.5) | SE | Partial | No | Yes |
| Venus-PowerX | 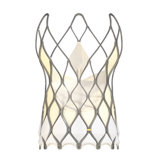 | 11 (1.4) | 0 (0) | SE | Full | Yes | Yes |
| TaurusOne | 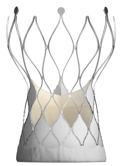 | 35 (4.4) | 0 (0) | SE | None | Yes | No |
| TaurusElite |  | 48 (6.1) | 0 (0) | SE | Full | Yes | Yes |
| TaurusNXT | 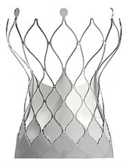 | 14 (1.8) | 0 (0) | SE | Full | Yes | Yes |
| VitaFlow | 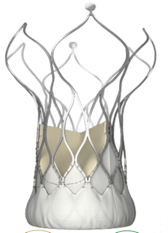 | 17 (2.1) | 0 (0) | SE | None | Yes | No |
| VitaFlow II |  | 58 (7.3) | 0 (0) | SE | Partial | Yes | Yes |
| Evolut PRO | 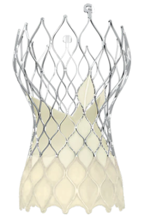 | 35 (4.4) | 0 (0) | SE | Full | Yes | Yes |
| ProStyle A | 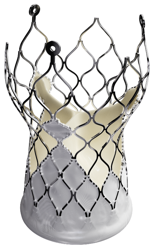 | 12 (1.5) | 0 (0) | SE | Full | Yes | Yes |
| SAPIEN XT | 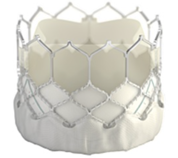 | 3 (0.4) | 0 (0) | BE | None | Yes | Yes |
| SAPIEN 3 |  | 11 (1.4) | 0 (0) | BE | None | Yes | Yes |
| LOTUS | 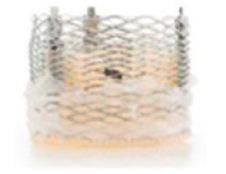 | 11 (1.4) | 0 (0) | ME | Full | Yes | Yes |
| Prizvalve | 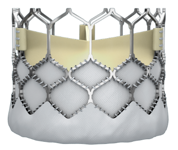 | 69 (8.7) | 0 (0) | BE | None | Yes | Yes |
| Prizvalve PRO |  | 2 (0.3) | 0 (0) | BE | None | Yes | Yes |
| Muguet A | 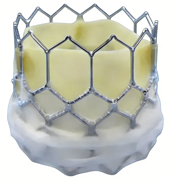 | 1 (0.1) | 0 (0) | BE | None | Yes | Yes |

Values are n (%).

### Table S2. Baseline Characteristics and Procedural Details of the Multicentre External Validation Cohort According to the BAV-Complex Score.

|  | All  N = 134 | Low risk  n= 65 | Intermediate risk  n= 65 | High risk n= 4 | p |
| --- | --- | --- | --- | --- | --- |
| Male | 85 (63.4) | 32 (49.2) | 49 (75.4) | 4 (100.0) | 0.002 |
| Age, yrs | 71 (66–74) | 70 (66–74) | 71 (66–75) | 66 (64–67) | 0.165 |
| BMI, kg/m² | 23.3 (22.2–24.2) | 23.3 (22.2–24.8) | 23.3 (22.0–23.6) | 23.4 (23.3–23.6) | 0.691 |
| STS score, % | 1.88 (1.34–3.33) | 1.87 (1.43–3.48) | 1.89 (1.24–2.85) | 1.84 (1.41–2.77) | 0.698 |
| Coronary artery disease | 11 (8.2) | 5 (7.7) | 6 (9.2) | 0 (0.0) | 1.000 |
| Chronic kidney disease | 5 (3.7) | 2 (3.1) | 3 (4.6) | 0 (0.0) | 1.000 |
| Atrial fibrillation | 8 (6.0) | 2 (3.1) | 6 (9.2) | 0 (0.0) | 0.434 |
| Aortic valve calcification volume, mm³ | 550 (288–1079) | 423 (168–711) | 797 (347–1185) | 1216 (1010–1430) | <0.001 |
| Aortic annular angle, ° | 52 (46–59) | 53 (46–60) | 51 (46–59) | 47 (44–53) | 0.620 |
| Aortic annular perimeter, mm | 77.6 (72.0–83.8) | 75.0 (69.4–78.8) | 80.4 (75.7–85.4) | 81.8 (79.9–86.0) | <0.001 |
| Aortic annular area, mm² | 471.2 (393.9–538.9) | 426.2 (369.6–490.6) | 498.5 (444.7–568.0) | 518.7 (494.6–579.6) | <0.001 |
| Sinus of Valsalva perimeter, mm | 106.0 (98.6–112.9) | 100.9 (93.7–106.0) | 109.1 (104.8–119.6) | 112.1 (111.2–114.8) | 0.002 |
| STJ diameter, mm | 31.1 (28.9–34.3) | 29.3 (27.6–31.4) | 33.5 (30.9–36.8) | 36.7 (35.8–38.8) | <0.001 |
| Maximum ascending aortic diameter, mm | 39.3 (36.1–42.8) | 37.4 (34.1–39.0) | 42.0 (39.6–44.0) | 47.8 (47.0–48.6) | <0.001 |
| LVOT perimeter, mm | 79.778 (72.534–87.920) | 74.104 (66.882–81.640) | 83.210 (77.558–91.374) | 89.702 (87.531–92.316) | <0.001 |
| Aortic regurgitation (≥ moderate) | 49 (36.6) | 25 (38.5) | 23 (35.4) | 1 (25.0) | 0.946 |
| Mean aortic valve gradient, mmHg | 54 (44–69) | 54 (45–68) | 54 (42–70) | 67 (54–80) | 0.679 |
| Peak aortic valve velocity, m/s | 4.65 (4.29–5.16) | 4.65 (4.40–5.15) | 4.64 (4.17–5.20) | 4.80 (4.56–5.13) | 0.596 |
| LVEF, % | 58 (46–61) | 59 (56–64) | 52 (39–60) | 36 (33–42) | <0.001 |
| LVEDD, mm | 50 (45–56) | 46 (43–50) | 54 (50–58) | 53 (51–57) | <0.001 |
| IVS, mm | 14 (12–15) | 13 (12–15) | 14 (12–15) | 14 (12–15) | 0.665 |
| Self-expandable THV | 134 (100.0) | 65 (100.0) | 65 (100.0) | 4 (100.0) | - |
| New generation THV | 133 (99.3) | 64 (98.5) | 65 (100.0) | 4 (100.0) | 1.000 |
| Pre-dilatation | 127 (94.8) | 62 (95.4) | 61 (93.8) | 4 (100.0) | 1.000 |
| Post-dilatation | 48 (35.8) | 25 (38.5) | 22 (33.8) | 1 (25.0) | 0.891 |

Values are n (%) and median (IQR).

BAV = bicuspid aortic valve; LVEDD = left ventricular end-diastolic diameter; LVEF = left ventricular ejection fraction; LVOT = left ventricular outflow tract; IVS =interventricular septum; STJ = sinotubular junction; STS = Society of Thoracic Surgeons.

**Table S3.** Procedural and Clinical Outcomes According to BAV-Complex Score Risk Groups.

|  | Development cohort | | | | | External validation cohort | | | | |
| --- | --- | --- | --- | --- | --- | --- | --- | --- | --- | --- |
|  | All  N = 793 | Low risk n = 276 | Intermediate risk  n = 445 | High risk n = 72 | p | All  N = 134 | Low risk  n = 65 | Intermediate risk  n = 65 | High risk  n = 4 | p |
| Intraprocedural death | 5 (0.6) | 1 (0.4) | 2 (0.4) | 2 (2.8) | 0.079 | 0 (0.0) | 0 (0.0) | 0 (0.0) | 0 (0.0) | - |
| Failure of device delivery | 2 (0.3) | 1 (0.4) | 1 (0.2) | 0 (0.0) | 1.000 | 0 (0.0) | 0 (0.0) | 0 (0.0) | 0 (0.0) | - |
| Implantation of multiple THVs | 47 (5.9) | 6 (2.2) | 27 (6.1) | 14 (19.4) | <0.001 | 1 (0.7) | 0 (0.0) | 1 (1.5) | 0 (0.0) | 1.000 |
| Conversion to open surgery | 1 (0.1) | 0 (0.0) | 1 (0.2) | 0 (0.0) | 1.000 | 1 (0.7) | 0 (0.0) | 0 (0.0) | 1 (25.0) | 0.030 |
| Cardiac tamponade | 6 (0.8) | 1 (0.4) | 3 (0.7) | 2 (2.8) | 0.170 | 1 (0.7) | 0 (0.0) | 1 (1.5) | 0 (0.0) | 1.000 |
| Aortic dissection | 7 (0.9) | 2 (0.7) | 4 (0.9) | 1 (1.4) | 0.712 | 2 (1.5) | 0 (0.0) | 2 (3.1) | 0 (0.0) | 0.526 |
| Coronary obstruction | 9 (1.1) | 4 (1.4) | 5 (1.1) | 0 (0.0) | 0.781 | 4 (3.0) | 2 (3.1) | 1 (1.5) | 1 (25.0) | 0.115 |
| Intraprocedural haemodynamic collapse requiring resuscitation | 35 (4.4) | 7 (2.5) | 21 (4.7) | 7 (9.7) | 0.027 | 2 (1.5) | 0 (0.0) | 2 (3.1) | 0 (0.0) | 0.526 |
| Major vascular complications | 16 (2.0) | 6 (2.2) | 9 (2.0) | 1 (1.4) | 1.000 | 3 (2.2) | 1 (1.5) | 2 (3.1) | 0 (0.0) | 1.000 |
| Major cardiac structural complications | 20 (2.5) | 4 (1.4) | 12 (2.7) | 4 (5.6) | 0.126 | 4 (3.0) | 2 (3.1) | 1 (1.5) | 1 (25.0) | 0.115 |
| Bleeding (Type 3 or 4) | 7 (0.9) | 1 (0.4) | 4 (0.9) | 2 (2.8) | 0.146 | 1 (0.7) | 0 (0.0) | 1 (1.5) | 0 (0.0) | 1.000 |
| Peak velocity at 30 days, m/s | 2.4 (2.0–2.7) | 2.4 (2.1–2.8) | 2.3 (2.0–2.7) | 2.4 (2.0–2.7) | 0.298 | 2.39 (2.02–2.63) | 2.39 (2.02–2.62) | 2.42 (2.03–2.70) | 2.30 (2.30–2.30) | 0.763 |
| Mean gradient at 30 days, mmHg | 12 (9–17) | 12 (9–18) | 12 (9–16) | 13 (9–16) | 0.561 | 13 (11–20) | 13 (11–19) | 15 (11–20) | 12 (12–12) | 0.941 |
| PVL≥moderate at 30 days | 13 (1.6) | 0 (0.0) | 8 (1.8) | 5 (6.9) | <0.001 | 9 (6.7) | 2 (3.1) | 6 (9.2) | 1 (25.0) | 0.108 |
| Acute kidney injury (Stage 3 or 4) | 4 (0.5) | 0 (0.0) | 4 (0.9) | 0 (0.0) | 0.404 | 1 (0.7) | 1 (1.5) | 0 (0.0) | 0 (0.0) | 1.000 |
| Covert CNS injury | 9 (1.1) | 4 (1.4) | 5 (1.1) | 0 (0.0) | 0.781 | 1 (0.7) | 0 (0.0) | 1 (1.5) | 0 (0.0) | 1.000 |
| Permanent pacemaker implantation | 124 (15.6) | 44 (15.9) | 61 (13.7) | 19 (26.4) | 0.023 | 5 (3.7) | 1 (1.5) | 4 (6.2) | 0 (0.0) | 0.456 |
| Related rehospitalization at 1 year | 12 (1.5) | 5 (1.8) | 5 (1.1) | 2 (2.8) | 0.314 | 4 (3.0) | 1 (1.5) | 2 (3.1) | 1 (25.0) | 0.115 |
| Clinical efficacy at 1 year | 722  (91.0) | 257  (93.1) | 403  (90.6) | 62  (86.1) | 0.155 | 127  (94.8) | 64  (98.5) | 60 (92.3) | 3 (75.0) | 0.062 |
| All-cause mortality at 1 year | 38 (4.8) | 7 (2.5) | 23 (5.2) | 8 (11.1) | 0.007 | 4 (3.0) | 1 (1.5) | 2 (3.1) | 1 (25.0) | 0.012 |
| Cardiovascular mortality at 1 year | 25 (3.2) | 5 (1.8) | 14 (3.1) | 6 (8.3) | 0.016 | 3 (2.2) | 1 (1.5) | 1 (1.5) | 1 (25.0) | 0.003 |

Values are n (%) and median (IQR).

### Table S4. Multivariable Analysis of the Composite Endpoint in the Development Cohort and External validation cohort

|  | OR (95% CI) | p |
| --- | --- | --- |
| **Development Cohort** | | |
| BAV-Complex score, per 1 point | 1.32 (1.18–1.47) | <0.001 |
| Age, per 1 year | 1.02 (0.99–1.06) | 0.113 |
| Male | 0.99 (0.62–1.61) | 0.972 |
| New-generation THV | 0.54 (0.34–0.83) | 0.005 |
| **External validation cohort*** | | |
| BAV-Complex score, per 1 point | 1.55 (1.09–2.32) | 0.021 |
| Age, per 1 year | 1.01 (0.91–1.12) | 0.892 |
| Male | 1.62 (0.35–11.48) | 0.568 |
| New-generation THV^a^ | - | - |

BAV = bicuspid aortic valve; CI = confidence interval; LVEF = left ventricular ejection fraction; OR =odds ratio; THV = transcatheter heart valve.

* In the external validation cohort, multivariable analysis was adjusted only for age and male sex due to the extremely high frequency of new-generation THV use (99.3%, 133/134).

### Table S5. Outcomes between Self-expandable and Balloon-expandable THVs in Development Cohort.

|  | Self-expandable THVs | | | | | Balloon-expandable/ Mechanically expandable THVs | | | | |
| --- | --- | --- | --- | --- | --- | --- | --- | --- | --- | --- |
|  | All  N = 696 | Low risk  n = 251 | Intermediate risk  n = 390 | High risk n = 55 | p | All  N = 97 | Low risk  n = 25 | Intermediate risk  n = 55 | High risk  n = 17 | p |
| **Composite endpoint at exit from procedure room** | 91 (13.1) | 18 (7.2) | 54 (13.8) | 19 (34.5) | <0.001 | 10 (10.3) | 2 (8.0) | 5 (9.1) | 3 (17.6) | 0.588 |
| Intraprocedural death | 3 (0.4) | 1 (0.4) | 2 (0.5) | 0 (0.0) | 1.000 | 2 (2.1) | 0 (0.0) | 0 (0.0) | 2 (11.8) | 0.029 |
| Failure of device delivery | 2 (0.3) | 1 (0.4) | 1 (0.3) | 0 (0.0) | 1.000 | 0 (0.0) | 0 (0.0) | 0 (0.0) | 0 (0.0) | - |
| Implantation of multiple THVs | 44 (6.3) | 5 (2.0) | 25 (6.4) | 14 (25.5) | <0.001 | 3 (3.1) | 1 (4.0) | 2 (3.6) | 0 (0.0) | 1.000 |
| Conversion to open surgery | 1 (0.1) | 0 (0.0) | 1 (0.3) | 0 (0.0) | 1.000 | 0 (0.0) | 0 (0.0) | 0 (0.0) | 0 (0.0) | - |
| Cardiac tamponade | 3 (0.4) | 1 (0.4) | 2 (0.5) | 0 (0.0) | 1.000 | 3 (3.1) | 0 (0.0) | 1 (1.8) | 2 (11.8) | 0.129 |
| Aortic dissection | 6 (0.9) | 2 (0.8) | 4 (1.0) | 0 (0.0) | 1.000 | 1 (1.0) | 0 (0.0) | 0 (0.0) | 1 (5.9) | 0.175 |
| Coronary obstruction | 8 (1.1) | 4 (1.6) | 4 (1.0) | 0 (0.0) | 0.855 | 1 (1.0) | 0 (0.0) | 1 (1.8) | 0 (0.0) | 1.000 |
| Intraprocedural haemodynamic collapse requiring resuscitation | 31 (4.5) | 6 (2.4) | 19 (4.9) | 6 (10.9) | 0.018 | 4 (4.1) | 1 (4.0) | 2 (3.6) | 1 (5.9) | 0.811 |
| Technical success at exit from procedure room | 622 (89.4) | 234 (93.2) | 349 (89.5) | 39 (70.9) | <0.001 | 88 (90.7) | 24 (96.0) | 50 (90.9) | 14 (82.4) | 0.309 |
| Major vascular complications | 14 (2.0) | 6 (2.4) | 8 (2.1) | 0 (0.0) | 0.700 | 2 (2.1) | 0 (0.0) | 1 (1.8) | 1 (5.9) | 0.386 |
| Major cardiac structural complications | 16 (2.3) | 4 (1.6) | 10 (2.6) | 2 (3.6) | 0.448 | 4 (4.1) | 0 (0.0) | 2 (3.6) | 2 (11.8) | 0.192 |
| Bleeding (Type 3 or 4) | 4 (0.6) | 1 (0.4) | 3 (0.8) | 0 (0.0) | 1.000 | 3 (3.1) | 0 (0.0) | 1 (1.8) | 2 (11.8) | 0.129 |
| Device success at 30 days | 485 (69.7) | 176 (70.1) | 281 (72.1) | 28 (50.9) | 0.006 | 74 (76.3) | 18 (72.0) | 44 (80.0) | 12 (70.6) | 0.613 |
| All-cause mortality at 30 days | 17 (2.4) | 3 (1.2) | 11 (2.8) | 3 (5.5) | 0.137 | 2 (2.1) | 0 (0.0) | 0 (0.0) | 2 (11.8) | 0.009 |
| Cardiovascular mortality at 30 days | 12 (1.7) | 3 (1.2) | 7 (1.8) | 2 (3.6) | 0.443 | 2 (2.1) | 0 (0.0) | 0 (0.0) | 2 (11.8) | 0.009 |
| Peak velocity at 30 days, m/s | 2.4 (2.0–2.7) | 2.4 (2.0–2.8) | 2.4 (2.0–2.7) | 2.4 (2.1–2.7) | 0.625 | 2.3 (2.1–2.5) | 2.4 (2.2–2.8) | 2.3 (2.1–2.5) | 2.1 (1.9–2.5) | 0.133 |
| Mean gradient at 30 days, mmHg | 12 (9–17) | 12 (9–18) | 12 (9–17) | 13 (10–16) | 0.570 | 11 (9–15) | 12 (10–17) | 11 (9–14) | 8 (7–15) | 0.087 |
| PVL≥moderate at 30 days | 13 (1.9) | 0 (0.0) | 8 (2.1) | 5 (9.1) | <0.001 | 0 (0.0) | 0 (0.0) | 0 (0.0) | 0 (0.0) | - |
| Early safety at 30 days | 537 (77.2) | 202 (80.5) | 301 (77.2) | 34 (61.8) | 0.012 | 78 (80.4) | 21 (84.0) | 48 (87.3) | 9 (52.9) | 0.011 |
| Acute kidney injury (Stage 3 or 4) | 4 (0.6) | 0 (0.0) | 4 (1.0) | 0 (0.0) | 0.288 | 0 (0.0) | 0 (0.0) | 0 (0.0) | 0 (0.0) | - |
| Covert CNS injury | 8 (1.1) | 3 (1.2) | 5 (1.3) | 0 (0.0) | 1.000 | 1 (1.0) | 1 (4.0) | 0 (0.0) | 0 (0.0) | 0.433 |
| Permanent pacemaker implantation | 111 (15.9) | 40 (15.9) | 57 (14.6) | 14 (25.5) | 0.121 | 13 (13.4) | 4 (16.0) | 4 (7.3) | 5 (29.4) | 0.051 |
| Clinical efficacy at 1 year | 633 (90.9) | 234 (93.2) | 351 (90.0) | 48 (87.3) | 0.233 | 89 (91.8) | 23 (92.0) | 52 (94.5) | 14 (82.4) | 0.270 |
| All-cause mortality at 1 year | 34 (4.9) | 7 (2.8) | 22 (5.6) | 5 (9.1) | 0.081 | 4 (4.1) | 0 (0.0) | 1 (1.8) | 3 (17.6) | 0.007 |
| Cardiovascular mortality at 1 year | 22 (3.2) | 5 (2.0) | 14 (3.6) | 3 (5.5) | 0.306 | 3 (3.1) | 0 (0.0) | 0 (0.0) | 3 (17.6) | <0.001 |
| Related rehospitalisation at 1 year | 12 (1.7) | 5 (2.0) | 5 (1.3) | 2 (3.6) | 0.290 | 0 (0.0) | 0 (0.0) | 0 (0.0) | 0 (0.0) | - |

Values are n (%) and median (IQR).

PVL = paravalvular leak ; THV = transcatheter heart valve.

### Table S6. Outcomes between First-generation and New-generation THVs in Development Cohort.

|  | First-generation THVs | | | | | New-generation THVs | | | | |
| --- | --- | --- | --- | --- | --- | --- | --- | --- | --- | --- |
|  | All  N = 389 | Low risk  n = 150 | Intermediate risk  n = 209 | High risk n = 30 | p | All  N = 404 | Low risk  n = 126 | Intermediate risk  n = 236 | High risk  n = 42 | p |
| Composite endpoint at exit from procedure room | 58 (14.9) | 11 (7.3) | 34 (16.3) | 13 (43.3) | <0.001 | 43 (10.6) | 9 (7.1) | 25 (10.6) | 9 (21.4) | 0.034 |
| Intraprocedural death | 2 (0.5) | 0 (0.0) | 2 (1.0) | 0 (0.0) | 0.585 | 3 (0.7) | 1 (0.8) | 0 (0.0) | 2 (4.8) | 0.011 |
| Failure of device delivery | 2 (0.5) | 1 (0.7) | 1 (0.5) | 0 (0.0) | 1.000 | 0 (0.0) | 0 (0.0) | 0 (0.0) | 0 (0.0) | - |
| Implantation of multiple THVs | 25 (6.4) | 2 (1.3) | 14 (6.7) | 9 (30.0) | <0.001 | 22 (5.4) | 4 (3.2) | 13 (5.5) | 5 (11.9) | 0.092 |
| Conversion to open surgery | 0 (0.0) | 0 (0.0) | 0 (0.0) | 0 (0.0) | - | 1 (0.2) | 0 (0.0) | 1 (0.4) | 0 (0.0) | 1.000 |
| Cardiac tamponade | 1 (0.3) | 0 (0.0) | 1 (0.5) | 0 (0.0) | 1.000 | 5 (1.2) | 1 (0.8) | 2 (0.8) | 2 (4.8) | 0.121 |
| Aortic dissection | 4 (1.0) | 0 (0.0) | 4 (1.9) | 0 (0.0) | 0.275 | 3 (0.7) | 2 (1.6) | 0 (0.0) | 1 (2.4) | 0.090 |
| Coronary obstruction | 8 (2.1) | 4 (2.7) | 4 (1.9) | 0 (0.0) | 0.856 | 1 (0.2) | 0 (0.0) | 1 (0.4) | 0 (0.0) | 1.000 |
| Intraprocedural haemodynamic collapse requiring resuscitation | 21 (5.4) | 3 (2.0) | 13 (6.2) | 5 (16.7) | 0.005 | 14 (3.5) | 4 (3.2) | 8 (3.4) | 2 (4.8) | 0.843 |
| Technical success at exit from procedure room | 342 (87.9) | 138 (92.0) | 185 (88.5) | 19 (63.3) | <0.001 | 368 (91.1) | 120 (95.2) | 214 (90.7) | 34 (81.0) | 0.018 |
| Major vascular complications | 10 (2.6) | 4 (2.7) | 6 (2.9) | 0 (0.0) | 1.000 | 6 (1.5) | 2 (1.6) | 3 (1.3) | 1 (2.4) | 0.703 |
| Major cardiac structural complications | 11 (2.8) | 3 (2.0) | 6 (2.9) | 2 (6.7) | 0.289 | 9 (2.2) | 1 (0.8) | 6 (2.5) | 2 (4.8) | 0.212 |
| Bleeding (Type 3 or 4) | 1 (0.3) | 0 (0.0) | 1 (0.5) | 0 (0.0) | 1.000 | 6 (1.5) | 1 (0.8) | 3 (1.3) | 2 (4.8) | 0.189 |
| Device success at 30 days | 254 (65.3) | 100 (66.7) | 141 (67.5) | 13 (43.3) | 0.031 | 305 (75.5) | 94 (74.6) | 184 (78.0) | 27 (64.3) | 0.158 |
| All-cause mortality at 30 days | 8 (2.1) | 1 (0.7) | 6 (2.9) | 1 (3.3) | 0.306 | 11 (2.7) | 2 (1.6) | 5 (2.1) | 4 (9.5) | 0.014 |
| Cardiovascular mortality at 30 days | 6 (1.5) | 1 (0.7) | 4 (1.9) | 1 (3.3) | 0.453 | 8 (2.0) | 2 (1.6) | 3 (1.3) | 3 (7.1) | 0.035 |
| Peak velocity at 30 days, m/s | 2.4 (2.1–2.8) | 2.5 (2.1–2.9) | 2.5 (2.1–2.8) | 2.3 (2.1–2.7) | 0.181 | 2.3 (1.9–2.6) | 2.2 (1.9–2.6) | 2.3 (1.9–2.6) | 2.4 (1.9–2.6) | 0.907 |
| Mean gradient at 30 days, mmHg | 13 (10–18) | 13 (10–18) | 13 (10–18) | 13 (10–18) | 0.584 | 11 (8–16) | 11 (8–16) | 11 (8–15) | 12 (8–16) | 0.806 |
| PVL≥moderate at 30 days | 10 (2.6) | 0 (0.0) | 6 (2.9) | 4 (13.3) | <0.001 | 3 (0.7) | 0 (0.0) | 2 (0.8) | 1 (2.4) | 0.197 |
| Early safety at 30 days | 286 (73.5) | 122 (81.3) | 149 (71.3) | 15 (50.0) | 0.001 | 329 (81.4) | 101 (80.2) | 200 (84.7) | 28 (66.7) | 0.019 |
| Acute kidney injury (Stage 3 or 4) | 1 (0.3) | 0 (0.0) | 1 (0.5) | 0 (0.0) | 1.000 | 3 (0.7) | 0 (0.0) | 3 (1.3) | 0 (0.0) | 0.680 |
| Covert CNS injury | 5 (1.3) | 2 (1.3) | 3 (1.4) | 0 (0.0) | 1.000 | 4 (1.0) | 2 (1.6) | 2 (0.8) | 0 (0.0) | 0.751 |
| Permanent pacemaker implantation | 72 (18.5) | 22 (14.7) | 39 (18.7) | 11 (36.7) | 0.018 | 52 (12.9) | 22 (17.5) | 22 (9.3) | 8 (19.0) | 0.040 |
| Clinical efficacy at 1 year | 355 (91.3) | 142 (94.7) | 186 (89.0) | 27 (90.0) | 0.158 | 367 (90.8) | 115 (91.3) | 217 (91.9) | 35 (83.3) | 0.200 |
| All-cause mortality at 1 year | 13 (3.3) | 1 (0.7) | 10 (4.8) | 2 (6.7) | 0.059 | 25 (6.2) | 6 (4.8) | 13 (5.5) | 6 (14.3) | 0.055 |
| Cardiovascular mortality at 1 year | 9 (2.3) | 1 (0.7) | 6 (2.9) | 2 (6.7) | 0.101 | 16 (4.0) | 4 (3.2) | 8 (3.4) | 4 (9.5) | 0.126 |
| Related rehospitalisation at 1 year | 11 (2.8) | 4 (2.7) | 5 (2.4) | 2 (6.7) | 0.370 | 1 (0.2) | 1 (0.8) | 0 (0.0) | 0 (0.0) | 0.416 |

Values are n (%) and median (IQR).

PVL = paravalvular leak ; THV = transcatheter heart valve.

### Table S7. Baseline, Anatomic, and Procedural Characteristics of Patients without Aortic Valve Calcification (n=12) versus the Rest of the Development Cohort (n=781).

|  | Zero calcium (n=12) | Other (n=781) | P |
| --- | --- | --- | --- |
| Age, years | 65 (64–70) | 72 (67–77) | 0.028 |
| Male sex | 5 (41.7%) | 451 (57.7%) | 0.001 |
| STS score, % | 1.9 (1.5–2.8) | 2.9 (1.9–4.8) | 0.080 |
| Coronary artery disease | 0 (0.0%) | 198 (25.4%) | 0.073 |
| Chronic kidney disease | 0 (0.0%) | 49 (6.3%) | 0.569 |
| Prior atrial fibrillation | 0 (0.0%) | 74 (9.5%) | 0.244 |
| Mean transvalvular gradient, mmHg | 44 (32–47) | 56 (44–71) | <0.001 |
| Peak aortic valve velocity, m/s | 4.2 (3.8–4.5) | 4.8 (4.3–5.4) | <0.001 |
| Left ventricular ejection fraction, % | 68 (66–73) | 61 (44–68) | <0.001 |
| Left ventricular end-diastolic diameter, mm | 49 (47–53) | 50 (45–56) | 0.050 |
| Interventricular septum thickness, mm | 12.0 (11.8–15.2) | 13.5 (12.0–15.0) | <0.001 |
| Moderate or greater aortic regurgitation | 2 (16.7%) | 104 (13.3%) | 0.885 |
| Moderate or greater mitral regurgitation | 1 (8.3%) | 93 (11.9%) | 0.885 |
| Moderate or greater tricuspid regurgitation | 0 (0.0%) | 49 (6.3%) | 0.202 |
| Aortic valve calcification volume, mm³ | 0 (0–0) | 517 (246–862) | <0.001 |
| Calcified raphe | 0 (0.0%) | 298 (38.2%) | <0.001 |
| Annular perimeter, mm | 75.2 (71.8–77.6) | 77.9 (71.9–84.3) | <0.001 |
| Annular area, mm² | 429 (400–462) | 467 (399–548) | <0.001 |
| LVOT perimeter, mm | 79.7 (74.8–82.5) | 82.1 (73.6–91.3) | <0.001 |
| Sinus of Valsalva perimeter, mm | 101.5 (97.8–105.7) | 109.1 (100.4–118.9) | <0.001 |
| Sinotubular junction diameter, mm | 28.6 (27.6–31.0) | 30.7 (28.1–33.7) | <0.001 |
| Maximal ascending aortic diameter, mm | 38.1 (37.0–41.9) | 42.0 (38.5–46.0) | <0.001 |
| Annulus angulation, ° | 52 (50–55) | 54 (48–61) | 0.704 |
| Self-expandable valve | 12 (100.0%) | 684 (87.6%) | 0.015 |
| New-generation THV | 8 (66.7%) | 392 (50.2%) | 0.108 |
| THV size, mm | 27 (26–29) | 26 (23–29) | 0.034 |
| Pre-dilatation | 8 (66.7%) | 734 (94.0%) | <0.001 |
| Post-dilatation | 3 (25.0%) | 411 (52.6%) | 0.033 |
| Contrast medium volume, mL | 300 (300–357) | 300 (270–350) | 0.104 |
| Composite intraprocedural endpoint | 1 (8.3%) | 100 (12.8%) | 0.802 |
| Implantation of multiple THVs | 1 (8.3%) | 46 (5.9%) | 0.342 |
| Valve migration | 1 (8.3%) | 14 (1.8%) | 0.115 |
| 30-day permanent pacemaker implantation | 5 (41.7%) | 119 (15.2%) | 0.043 |
| Moderate paravalvular leak at 30 days | 0 (0.0%) | 13 (1.7%) | 0.850 |
| 30-day all-cause mortality | 0 (0.0%) | 19 (2.4%) | 0.861 |
| 1-year all-cause mortality | 1 (8.3%) | 37 (4.7%) | 0.744 |

Values are n (%) and median (IQR).

Values shown for the zero-calcium subgroup (n=12) and the rest of the development cohort (n=781). All P values are 3-tier comparisons across the calcium strata (zero, 1–100, >100 mm³): Kruskal–Wallis test for continuous variables (median, IQR) and Pearson chi-square test for categorical variables (n, %). LVEF = left ventricular ejection fraction; LVOT = left ventricular outflow tract; STS = Society of Thoracic Surgeons; THV = transcatheter heart valve.

### Figure S1. Study Flowchart.

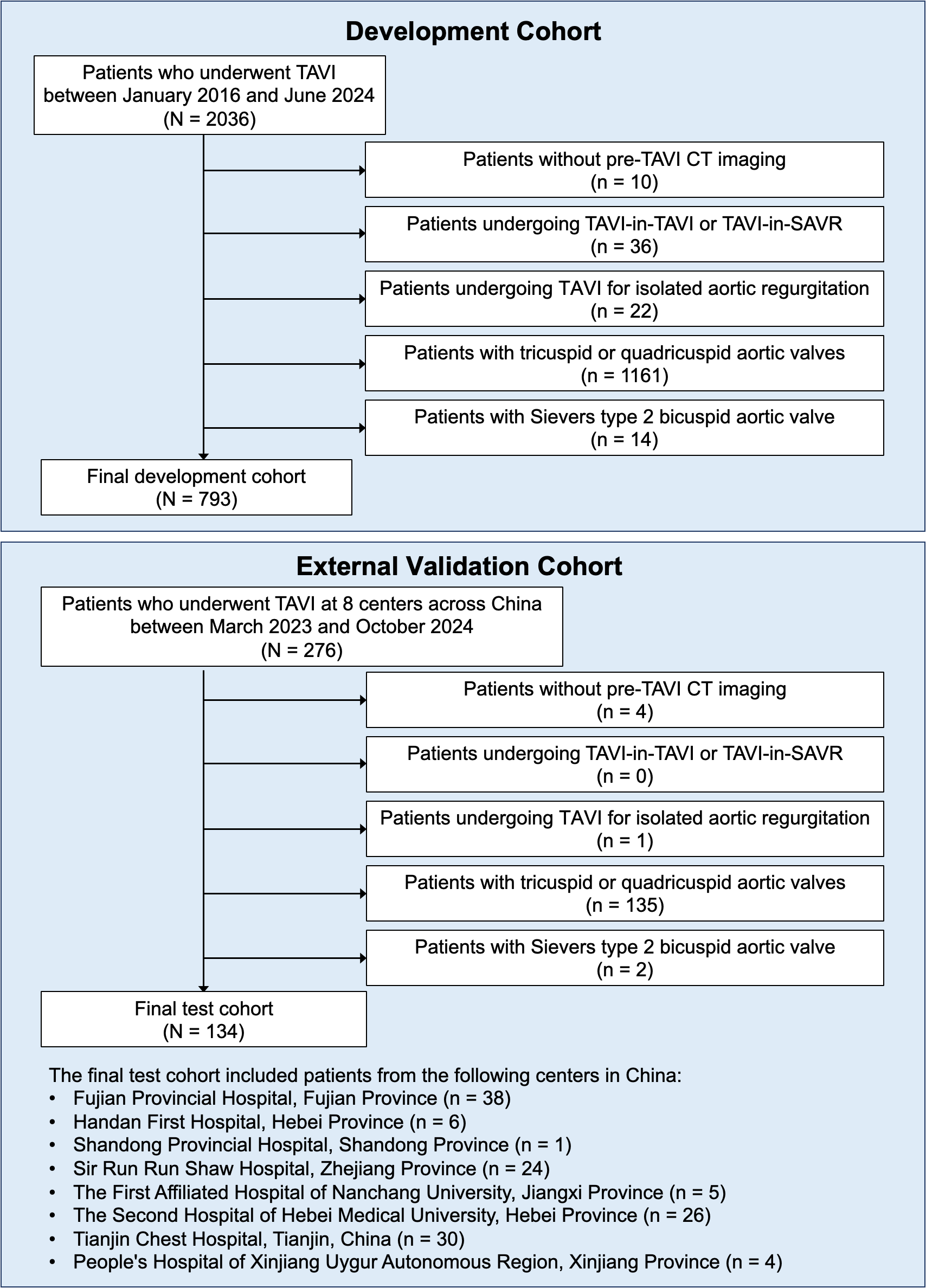

BAV = bicuspid aortic valve; TAVI = transcatheter aortic valve implantation; SAVR = surgical aortic valve replacement

### Figure S2. Incidence of the Composite Endpoint by Individual Anatomical Parameters in the Development Cohort.

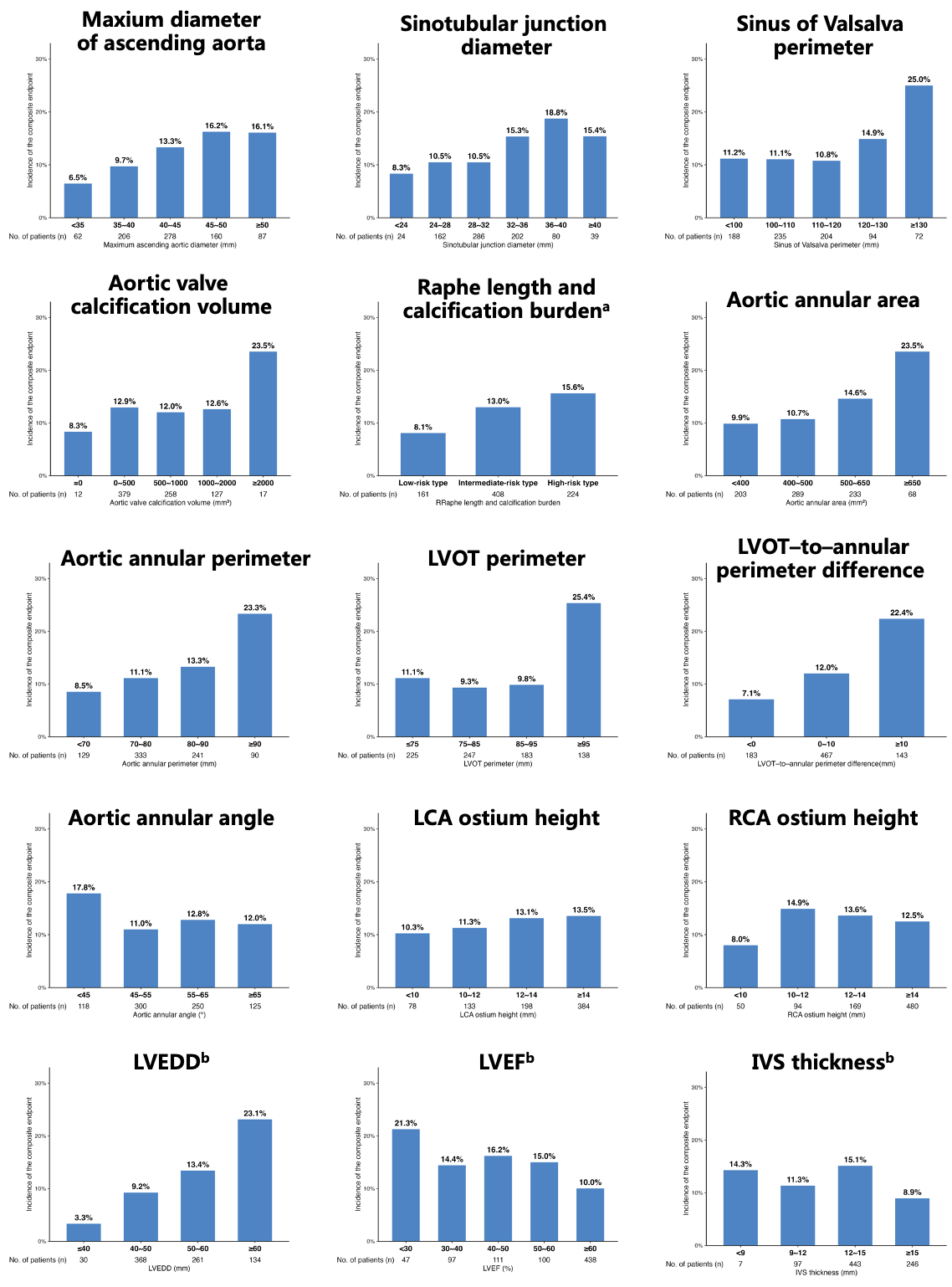

^a^ Raphe length and calcification burden refer to: Low-risk type= non-calcified raphe or calcified raphe ≤ 1/3 of the commissure edge; intermediate-risk type = no raphe; high-risk type= calcified raphe > 1/3 of the commissure edge.

^b^ Measured from transthoracic echocardiography.

IVS = interventricular septum; LCA = left coronary artery; LVEDD = left ventricular end-diastolic diameter; LVEF = left ventricular ejection fraction; LVOT = left ventricular outflow tract; RCA = right coronary artery.

### Figure S3. Anatomical variable screening, stability assessment, and contribution analysis for model development.

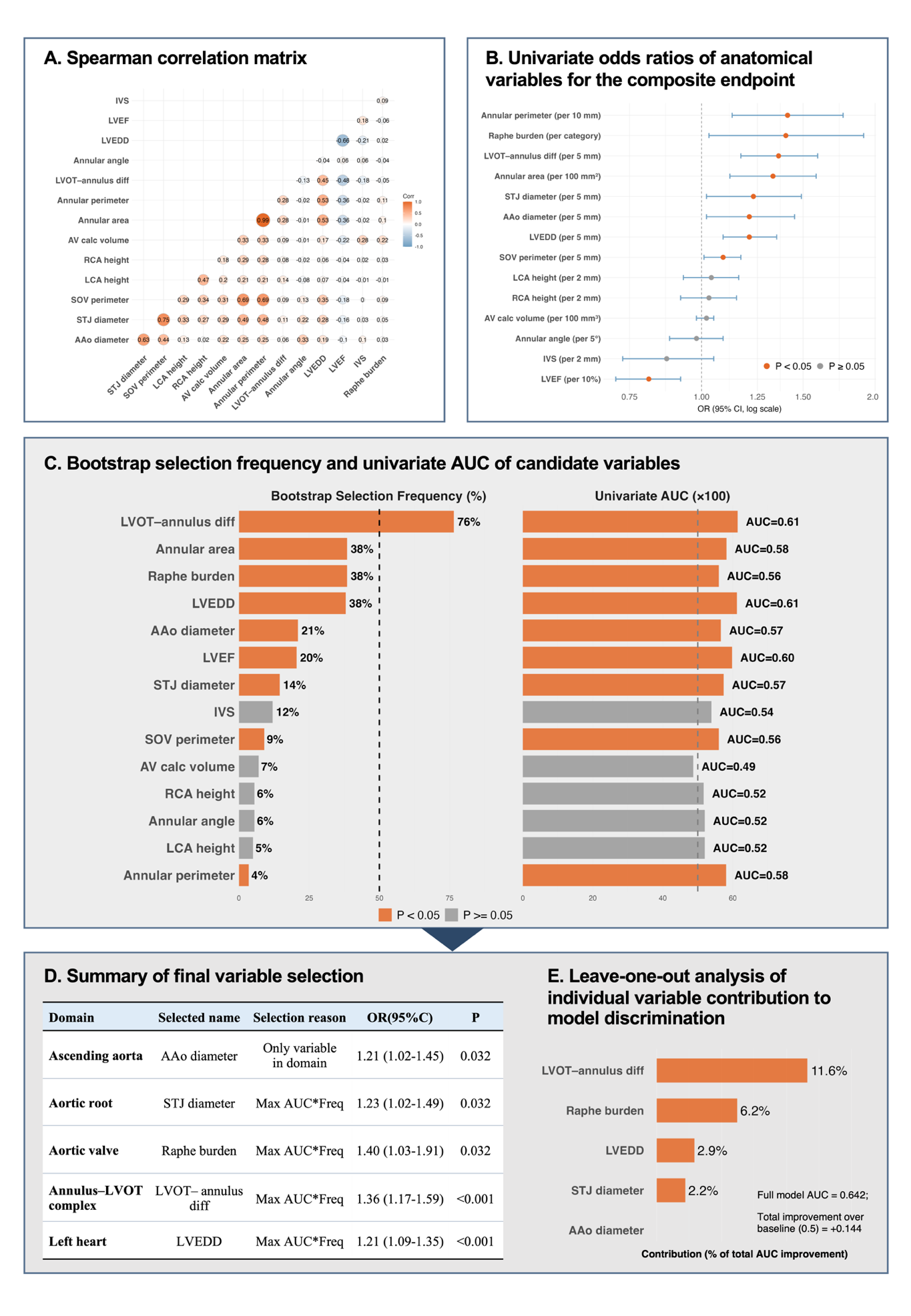

**(A)** Spearman correlation matrix illustrating collinearity among candidate anatomical indicators.
**(B)** Univariate logistic regression analysis showing odds ratios (ORs) and 95% confidence intervals for the association between each anatomical variable and the composite endpoint.
**(C)** Bootstrap-based stability analysis and univariate discriminative performance of candidate variables, quantified by selection frequency and area under the receiver operating characteristic curve (AUC).
**(D)** Final domain-constrained selection of anatomical variables retained for model construction.
**(E)** Leave-one-out analysis demonstrating the relative contribution of individual variables to model discrimination, calculated as the proportion () of total AUC improvement beyond the baseline value of 0.5.

**Abbreviations used in this figure:**
AAo diameter = maximum ascending aortic diameter; STJ diameter = sinotubular junction diameter; SOV perimeter = sinus of Valsalva perimeter; LCA height = left coronary artery ostium height; RCA height = right coronary artery ostium height; AV calc volume = aortic valve calcification volume; Annular area = aortic annular area; Annular perimeter = aortic annular perimeter; LVOT–annulus diff = LVOT–to–annular perimeter difference; Annular angle = aortic annular angulation; LVEDD = left ventricular end-diastolic diameter; LVEF = left ventricular ejection fraction; IVS = interventricular septal thickness.

### Figure S4. Group-Level External Calibration Before and After Recalibration.

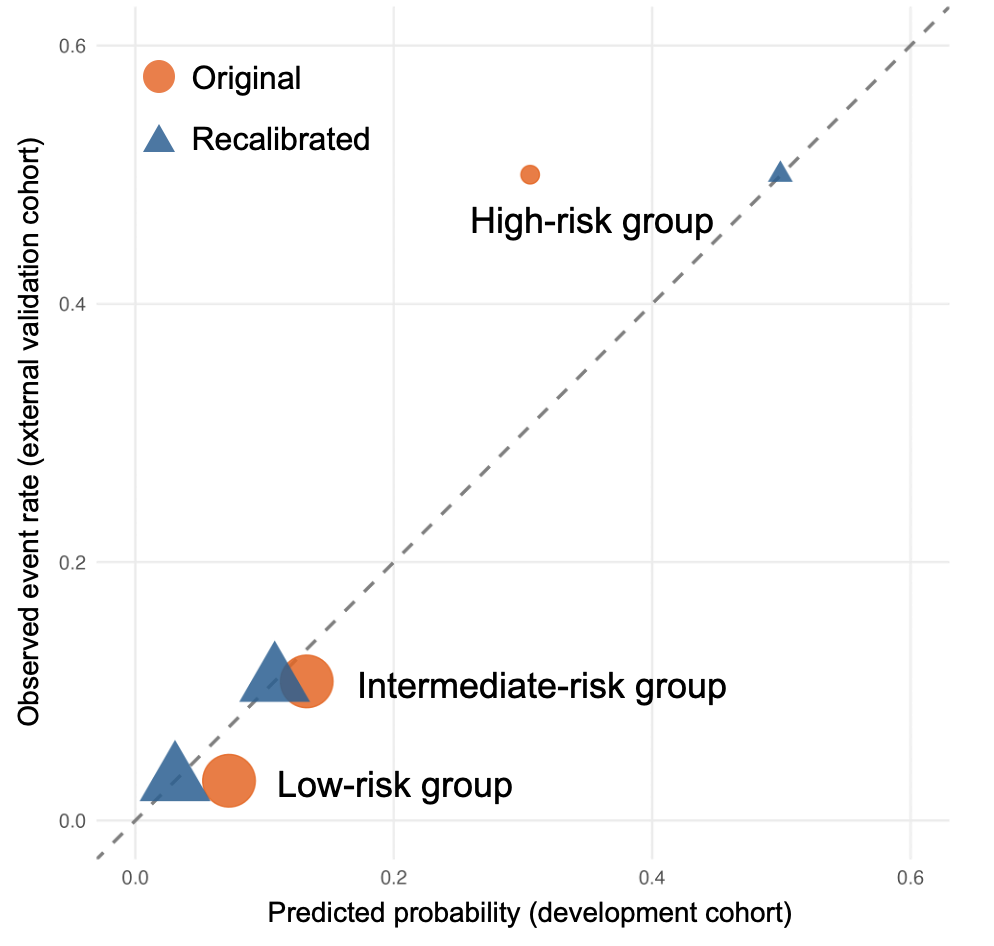

Observed event rates in the external validation cohort are plotted against predicted probabilities derived from the development-cohort group model for low- (score 0–3), intermediate- (4–7), and high-risk (8–10) categories. The dashed diagonal line represents perfect calibration. Point size is proportional to the number of patients per stratum. Predicted risk overestimated observed risk in the low-risk (7.2% vs 3.1%; n=65) and intermediate-risk strata (13.3% vs 10.8%; n=65), whereas it underestimated observed risk in the high-risk stratum (30.6% vs 50.0%; n=4). Calibration-in-the-large was −0.315 (95% CI, −0.996 to 0.260), and the calibration slope was 1.996 (95% CI, 0.641 to 3.516). Estimates for the high-risk stratum should be interpreted with caution given the limited sample size (n=4).

### Figure S5. Procedural and Outcome Comparison Across Aortic Valve Calcium Volume Strata.

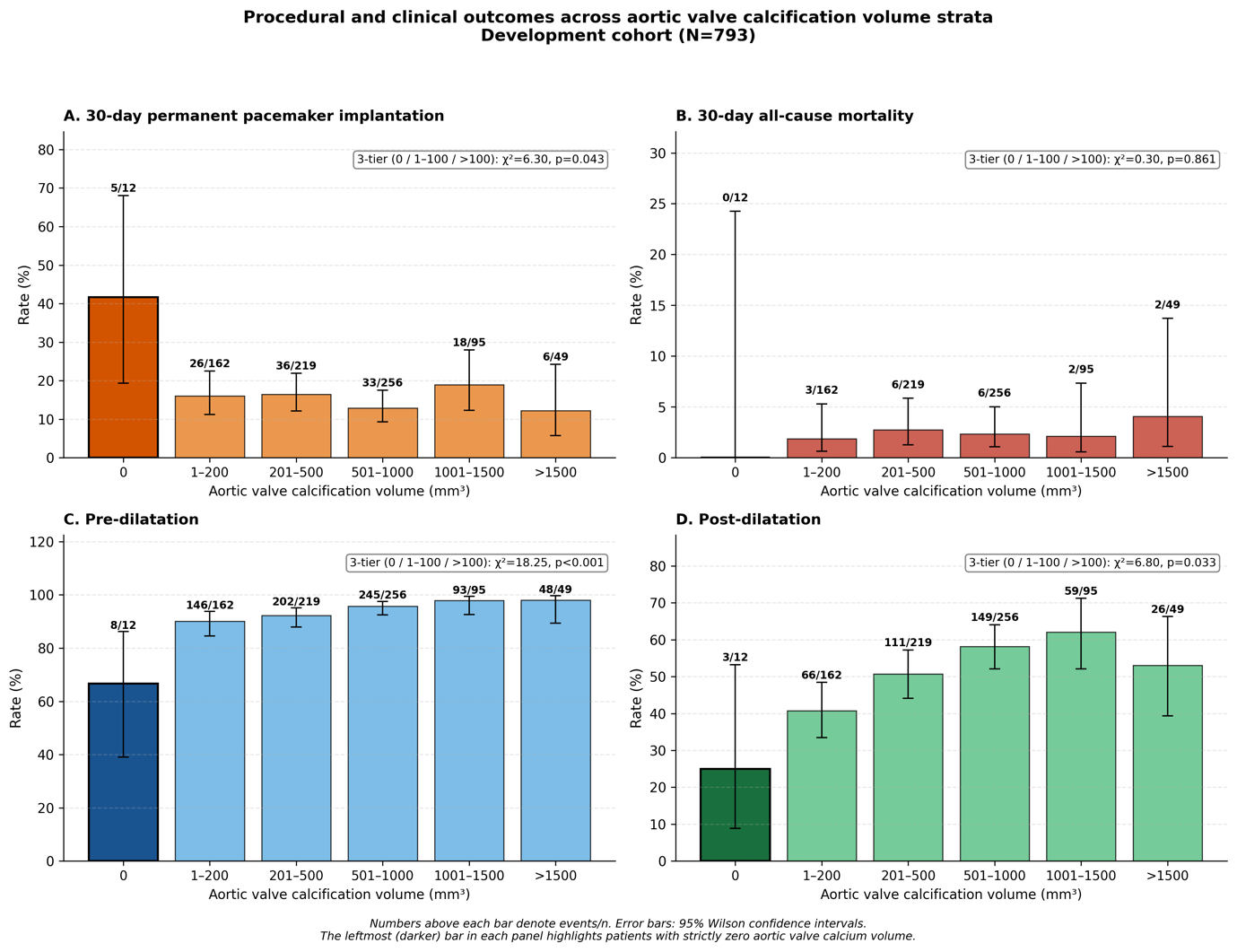
Four-panel comparison of patients with absent (zero, n=12), low (1–100 mm³, n=83), and higher (>100 mm³) aortic valve calcification in the development cohort. **(A)** 30-day permanent pacemaker implantation rate across calcium strata. **(B)** 30-day all-cause mortality across calcium strata. **(C)** Pre-dilatation utilisation across calcium strata. **(D)** Post-dilatation utilisation across calcium strata. P values from 3-tier chi-square test.
